# Predicting the Effect of Malaria Control Strategies Using Mathematical Modeling Approach

**DOI:** 10.1101/2020.10.28.20221267

**Authors:** Felix Yakubu Eguda, Samuel Eneojo Abah, James Andrawus, Sunday Atuba, Aliyu Abba

**Affiliations:** Department of Mathematics, Federal University Dutse, Nigeria; Department of Applied Biology, Coventry University, United Kingdom; British Community Hospital and Outpatient Clinics Ipaja, Lagos, Nigeria; British Pathodiagnostic and Biotechnology Laboratory, Ipaja, Lagos, Nigeria; Department of Engineering, Coventry University, United Kingdom; Department of Statistics, Jigawa State Polytechnic, Dutse, Nigeria

**Keywords:** Malaria, Mathematical model, Bifurcation, Stability

## Abstract

Malaria is a life-threatening disease which has caused enormous public health challenge. A mathematical model describing the dynamics of malaria between the human and vector population is formulated to understand the important parameters in the transmission and develop effective prevention and control strategies. We analysed the model and found that the model has a disease-free equilibrium (DFE) which is locally and globally asymptotically stable if the effective reproduction number can be brought below unity. Our model shows that the infectivity of mildly infected children and adults amplifies the disease burden in a population. It was shown that the model does not undergo the phenomenon of backward bifurcation so long as the recovered children and adults do not lose their acquired immunity and if the infection of mildly infected adult is not high enough to infect susceptible mosquitoes. However, control strategies involving mosquito reduction through high rate of application of insecticide will serve as an effective malaria control strategy. It is further shown that whenever the effective reproduction number is greater than unity the model has a unique endemic equilibrium which is globally stable for the case when there is loss of acquired immunity in children and adults. Numerical simulations show that the presence of all the control strategies is more effective in preventing mild malaria cases in adult and children as compared to severe malaria cases in adult and children.

## 1 Introduction

Malaria poses a substantial public health problem with about 228 million cases worldwide and 405,000 deaths globally [1]. Majority of this malaria burden up to 93% occur in Africa of which 85% is within the sub-Saharan Africa, of which Nigeria bears 25% of this burden. Hence, the country with the highest malaria burden [1]. Children under 5 years of age are the most affected and an increasing re-emergence of some severe clinical manifestation in adults is likely [1–3]. Malaria is a life-threatening disease that is transmitted through the bites of infected female anopheles mosquito (vector). After entering a human, the parasites transform through a complicated life-cycle in the liver and bloodstream. A stage in the life cycle developed into gametocytes, which spreads through a susceptible mosquito that bites the infectious human [3]. After approximately 10 to 15 days the mosquito takes her next blood meal and can infect a new person. After a human gets bitten, the symptoms appear in about 9-14 days. Clinical symptoms such as fever, pain, chills and sweats may develop a few days after infected mosquito bites. The infection can lead to serious complications affecting the brain, lungs, kidneys and other organs [3–5]. Since malaria increases morbidity and mortality, it continues to inflict major public health and socioeconomic burdens in developing countries. Malaria control even in countries with relatively low malaria endemicity proves to be a significant challenge. The complexity of the disease control process, the cost of the control programme and resistance of the parasite to anti-malarial drugs, and vectors to insecticides, are some of the challenges [3, 4, 6]. The rate of acquisition of immunity to severe malaria depends on age distribution of humans and the level of exposure to infections [7, 8]. Recently, the international community has increased its focus on eradicating malaria burden worldwide [1].

The battle towards the eradication and or control of malaria would have to take a collaborative approach to be achieved. Partly involving the role of mathematicians and their modelling approach in studying the dynamics of malaria, giving an insight into the interaction between the host and vector population and how to control its transmission. This is a collaborative work aimed at constructing a vector-borne compartmental model in a heterogeneous population incorporating mosquito reduction strategy, personal protection strategy, vaccination as control strategies and immune compartments in both children and adult population. This work considers all of these control strategies, which were not completely captured in previous report [8–10].

## 2 Model Formulation

A mathematical model for endemic malaria is formulated in a heterogeneous population with two human populations consisting of adults and children. The vector (mosquito) population is considered in this work where *N*_*A*_(*t*), *N*_*C*_(*t*) and *N*_*V*_ (*t*) denote the total number of adults, children and vectors at time *t*, respectively. As specified in (**Table 2.1**), the total population of human and vectors is divided into the following mutually exclusive epidemiological classes, namely, susceptible adults (*S*_*A*_(*t*)), adults with asymptomatic malaria (*E*_*A*_(*t*)), adults with malaria at mild stage (*I*_*AM*_ (*t*)), adults with severe malaria (*I*_*AS*_(*t*)), adults treated of malaria (*R*_*A*_(*t*)), immune adults (*R*(*t*)), susceptible children (*S*_*C*_(*t*)), children with asymptomatic malaria (*E*_*C*_(*t*)), children with mild malaria (*I*_*CM*_ (*t*)),children with severe malaria (*I*_*CS*_(*t*)),children treated of malaria (*R*_*C*_(*t*)), immune children (*Q*(*t*)), susceptible vectors (*S*_*V*_ (*t*)), vectors with parasite at latent stage(*E*_*V*_ (*t*)), vectors with parasite(*I*_*V*_ (*t*)), Hence, we have that in,

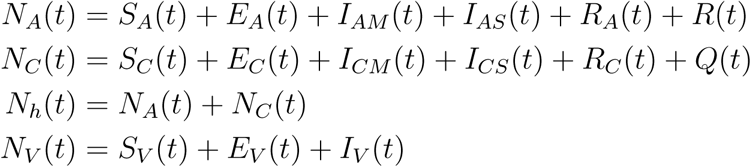

**Figure A:**
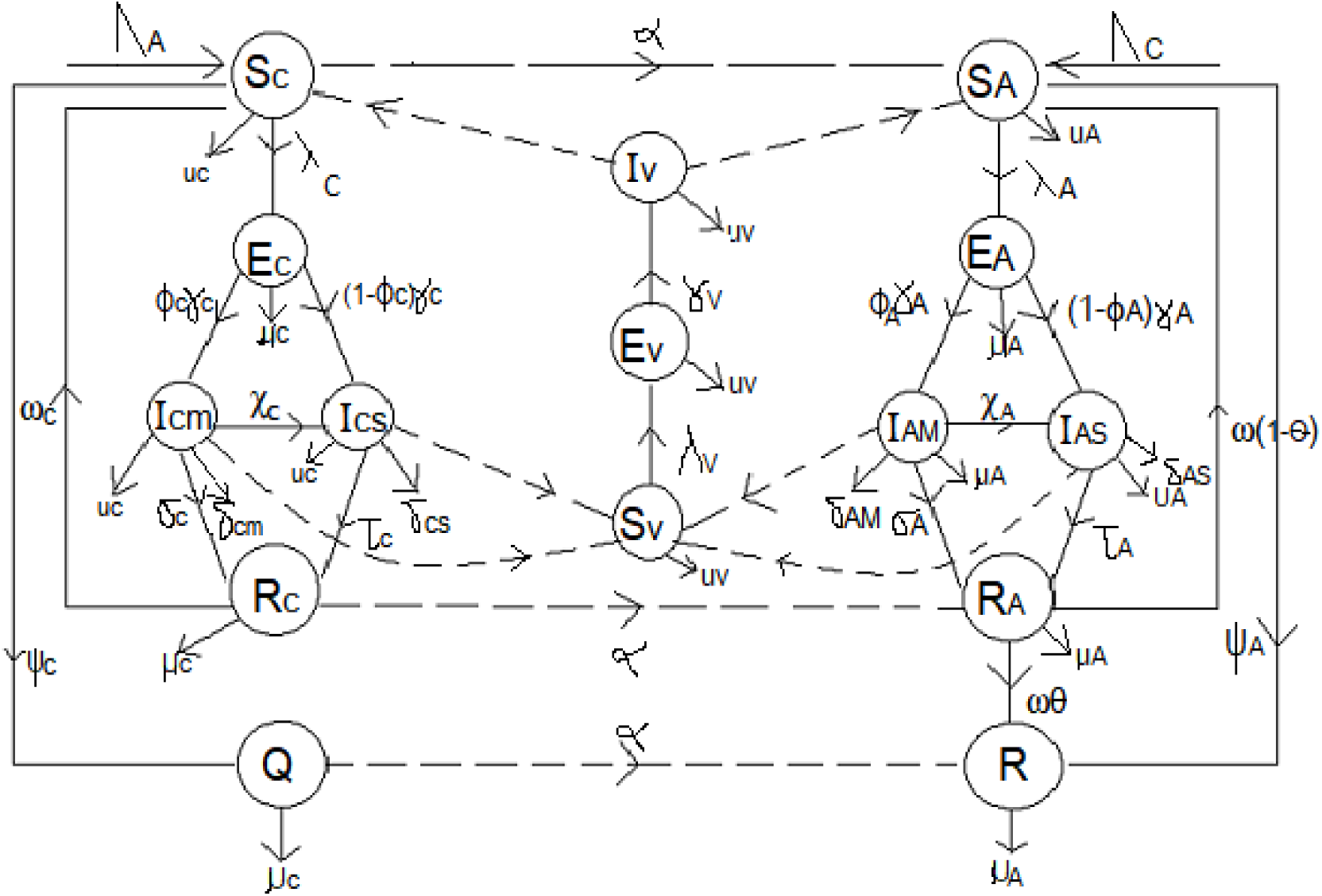
Flow diagram of malaria model

Susceptible adults contact malaria with disease force of infection rate

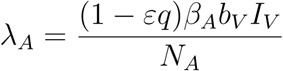

Susceptible children contact malaria at a rate

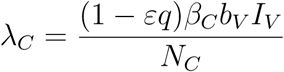

Susceptible vectors acquire the gametocytes from infected humans at a rate

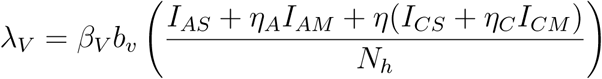

Susceptible children and adults are recruited at rates Λ_*C*_, Λ_*A*_ while the susceptible mosquitoes are recruited at a rate Λ_*V*_ (**Table 2.1**). Also, susceptible adults and children are vaccinated at rates *ψ*_*A*_, *ψ*_*C*_ respectively. We assume that fraction *ϕ*_*A*_, *ϕ*_*C*_ of adults and children with asymptomatic malaria progress to the mild stage of infection (at rates *γ*_*A*_, *γ*_*C*_ respectively) while remaining fraction 1 − *ϕ*_*A*_, 1− *ϕ*_*c*_ of adults and children progress to the severe stage of infection (at rates *γ*_*A*_, *γ*_*C*_).

Let *β*_*A*_, *β*_*C*_, *β*_*V*_ represent transmission probability per contact for adults, children and susceptible mosquitoes. Infected adults at mild and severe stages of infection progress to class of recovered adults at rates *σ*_*A*_, *τ*_*A*_ respectively, while infected children at mild and severe stages of infection progress to class or recovered children at rates, *σ*_*C*_, *τ*_*C*_ respectively.

The parameters *δ*_*AM*_, *δ*_*AS*_, *δ*_*CM*_, *δ*_*CS*_, represent the disease induced death rates for adults and children at mild and severe stage respectively. *δ*_*V*_ represent the death rate of mosquitoes from insecticide. The parameters *θ* represent the fraction of recovered adults who develop immunity after recovery while 1 − *θ* represent the remaining fraction of recovered adults who become susceptible after treatment. *b*_*V*_ is the mosquito biting rate while *q* is the rate of insecticide treated nets (ITN) compliance, *ω* represent the rate at which recovered adults revert to either the immune or susceptible adult class while *ω*_*C*_ is the rate at which recovered children revert to susceptible children class. Vectors at latent stage become infectious at the rate *γ*_*V*_. The parameters *η*_*A*_, *η*_*C*_ represent the infectivity modification parameters in adults and children. *η* is a modification parameter that indicates that children exposure rate is different from that of adults. That is, children protected from mosquito bites are less likely to get malaria infections. We assume that the infection in the mild classes might not be high enough to infect susceptible mosquitoes or is at the same level as the infectious individual giving 0 ≤ *η*_*C*_, *η*_*A*_ ≤ 1.

The model equations are given below

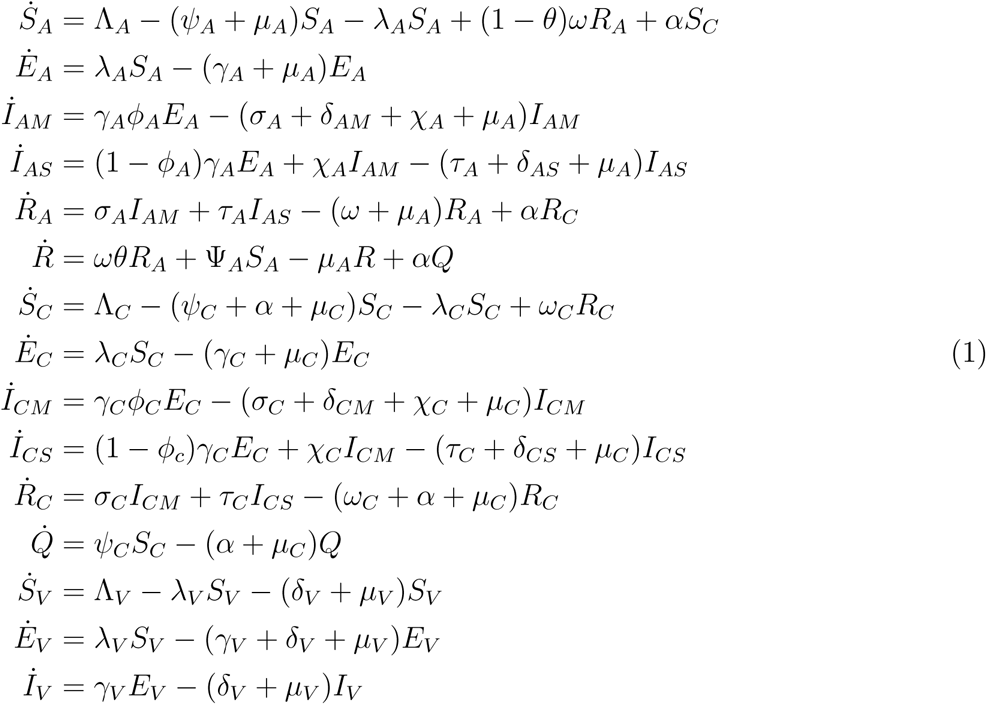

### 2.1 Basic Properties

Since the model monitors both human and mosquito population during malaria epidemic, it is important to prove that all the state variables of the model are non-negative for all time (*t*) for the model (1) to be epidemiologically meaningful. That is, the solutions of the model (1) with positive initial data will remain positive for all time *t* > 0. Model (1) is basically divided into two regions, thus *D* = *D*_1_ × *D*_2_.

#### 2.1.1 Boundedness

##### Lemma 1.

*The region D* = *D*_1_ × *D*_2_. *of system (1) is positively invariant with non-negative initial conditions in* 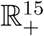

***Proof:*** The rate of change of the total human population is given as

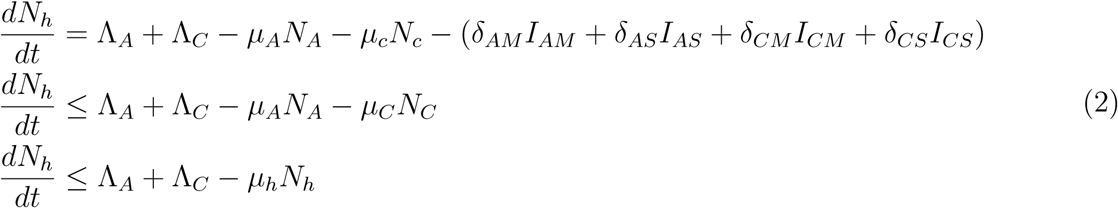

**Table 2.1:** Description of the variables and parameters of the Malaria model

| Variable | Description |
| --- | --- |
| $S_A, S_C$ | Population of susceptible adults and children |
| $E_A, E_C$ | Population of adults and children with asymptomatic malaria |
| $I_{AM}, I_{CM}$ | Population of infected adults and children at mild stage |
| $I_{AS}, I_{CS}$ | Population of infected adults and children at severe stage |
| $R_A, R_C$ | Population of recovered adults and children |
| $R, Q$ | Immune adults and children |
| $S_V, E_V, I_V$ | Susceptible, Exposed and Infected Mosquitoes |
| Parameter | Description |
| $\Lambda_A, \Lambda_C$ | Recruitment rate into the population of adults and children respectively |
| $\Lambda_V$ | Vector recruitment rate |
| $\mu_A, \mu_C, \mu_C$ | Natural death rate for humans (adults & children), mosquitoes respectively |
| $\beta_A, \beta_C, \beta_V$ | Transmission probability per contact for adults, children and susceptible mosquitoes |
| $\gamma_A, \gamma_C,$ | Rate at which exposed adults and children revert to the mild stage of infection |
| $\phi_A, \phi_C$ | Fraction of exposed adults and children who progress to the mild stage of infection |
| $\delta_{AM}, \delta_{AS}$ | Disease induced death rates of adults at mild and severe stage |
| $\delta_{CM}, \delta_{CS}$ | Disease induced death rates of children at mild and severe stage |
| $\delta_V$ | Death rate of mosquitoes from insecticide |
| $\sigma_A, \tau_A$ | Treatment rates for adults at mild and severe stage |
| $\sigma_C, \tau_C$ | Treatment rates for children at mild and severe stage |
| $\theta$ | Fraction of recovered adults who develop immunity after recovery |
| $\omega$ | Rate at which recovered adults revert to either the immune or susceptible adult class |
| $\omega\theta$ | Acquisition of immunity in adults |
| $\omega(1 - \theta)$ | Loss of immunity in adults |
| $\alpha$ | Growth and maturation rate |
| $\psi_A, \psi_C$ | Vaccination rate for adults and children |
| $\omega_C$ | Rate at which recovered children revert to susceptible children class |
| $\gamma_V$ | Rate at which exposed mosquitoes become infectious |
| $b_V$ | Mosquito biting rate |
| $\varepsilon, 0 \leq \varepsilon \leq 1$ | Efficacy of insecticide treated nets(ITN) |
| $q$ | Rate of ITN compliance |
| $\eta$ | modification parameter |
| $\eta_A, \eta_C$ | Infectivity modification parameters in adults and children |
| $\chi_A, \chi_C$ | Rate at which infected adults and children at mild stage progress to the severe stage of infection |

Where *μ*_*h*_ = min {*μ*_*A*_, *μ*_*C*_}

A standard comparison theorem [11] can then be used to show that

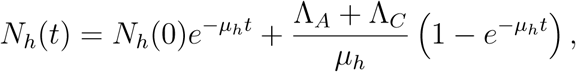

In particular, if

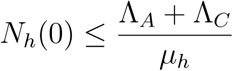

then,

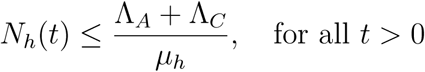

The rate of change of the total vector population is given as

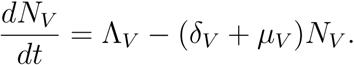

By standard comparison theorem,

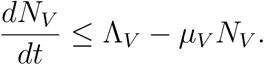

Solving gives

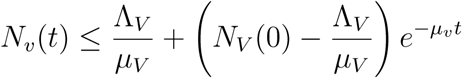

In particular, if

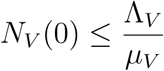

then,

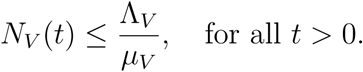

Then either the solution enters *D* in finite time or *N*_*V*_ (*t*) approaches 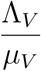 as *t* → ∞. Thus, the region *D* = *D*_1_ × *D*_2_ is positively invariant so that no solution path leaves through any boundary of *D*. Thus the feasible solution of the human population is in the region

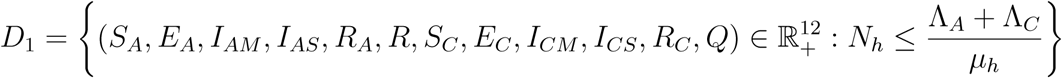

and the feasible solution of the vector population is in the region

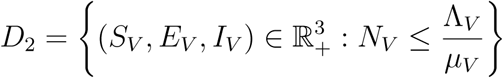

Hence it is sufficient to consider the dynamics of the model (1) in the region. In this region, the model can be considered as being mathematically and epidemiologically well posed. Hence, the solution set of model (1) are contained in *D*.

### 2.2 Positivity of Solutions

We assumed that the initial conditions of the model are non-negative and we also showed that the solution of the model is also positive.

#### Lemma 2.

*Let the initial data for the model be*

*S*_*A*_(0) > 0, *E*_*A*_(0) > 0, *I*_*AM*_ (0) > 0, *I*_*AS*_(0) > 0, *R*_*A*_(0) > 0, *R*(0) > 0, *S*_*C*_(0) > 0, *E*_*C*_(0) > 0, *I*_*CM*_ (0) > 0, *I*_*CS*_(0) > 0, *R*_*C*_(0) > 0, *Q*(0) > 0, *S*_*V*_ (0) > 0, *E*_*V*_ (0) > 0, *I*_*V*_ (0) > 0, *then the solutions* (*S*_*A*_, *E*_*A*_, *I*_*AM*_, *I*_*AS*_, *R*_*A*_, *R, S*_*C*_, *E*_*C*_, *I*_*CM*_, *I*_*CS*_, *R*_*C*_, *Q, S*_*V*_, *E*_*V*_, *I*_*V*_) *of the model (1) with initial data will remain positive for all time t* > 0.

***Proof:***

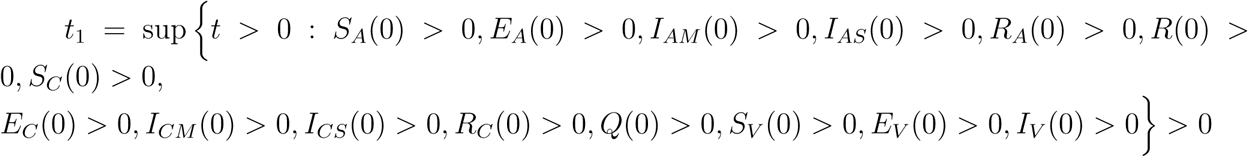

From the first equation in model (1)

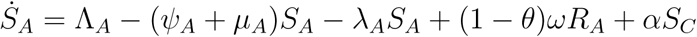

which implies

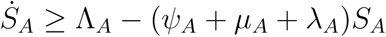

Using integrating factor method, we have

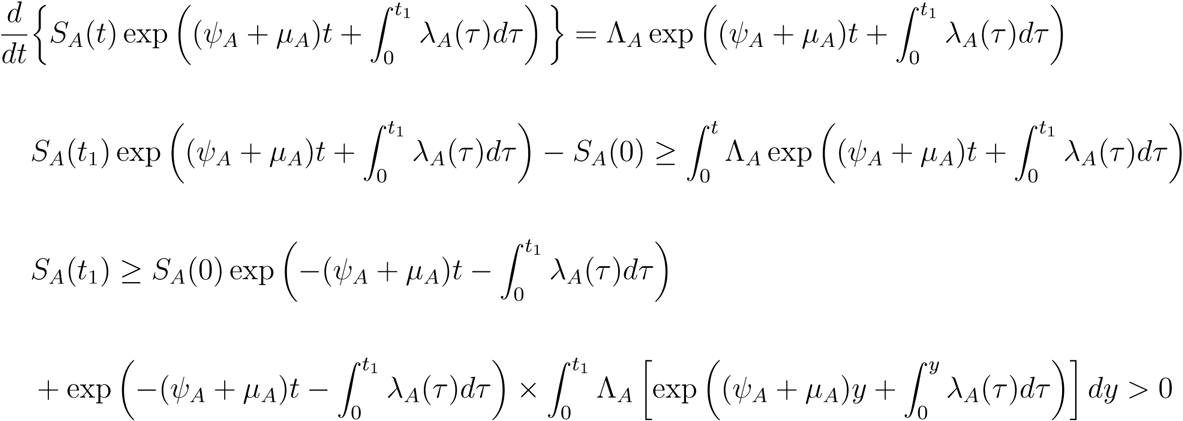

Similarly, it can be shown that all state variables of the model remain positive for all time *t* > 0 *E*_*A*_(*t*) > 0, *I*_*AM*_ (*t*) > 0, *I*_*AS*_(*t*) > 0, *R*_*A*_(*t*) > 0, *R*(*t*) > 0, *S*_*C*_(*t*) > 0, *E*_*C*_(*t*) > 0, *I*_*CM*_ (*t*) > 0, *I*_*CS*_(*t*) > 0, *R*_*C*_(*t*) > 0, *Q*(*t*) > 0, *S*_*V*_ (*t*) > 0, *E*_*V*_ (*t*) > 0, *I*_*V*_ (*t*) > 0 for all time *t* > 0.

## 3 Analysis of the Model

### 3.1 Local stability of the disease-free equilibrium(DFE)

The disease-free equilibrium of the model (1) is given by

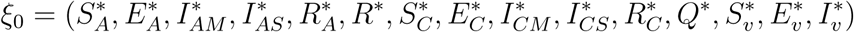

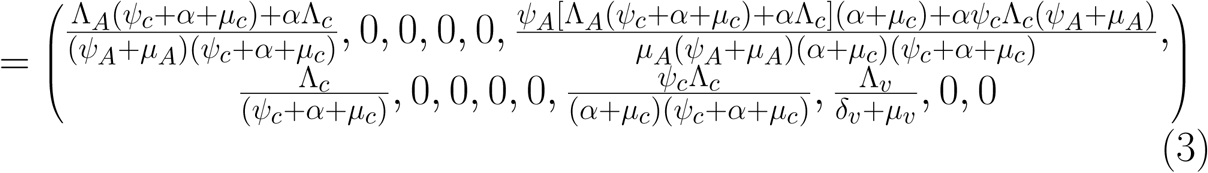

Using the next generation operator method previously described [12],the local stability of *ξ*_0_ can be established, it follows that matrices, *F* and *V* for the new infection terms and the remaining transition terms are respectively given by

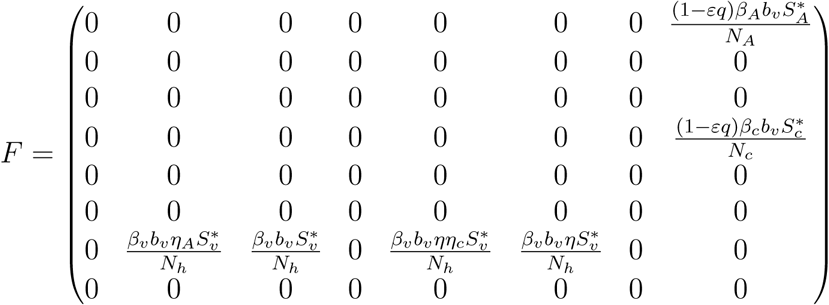

and

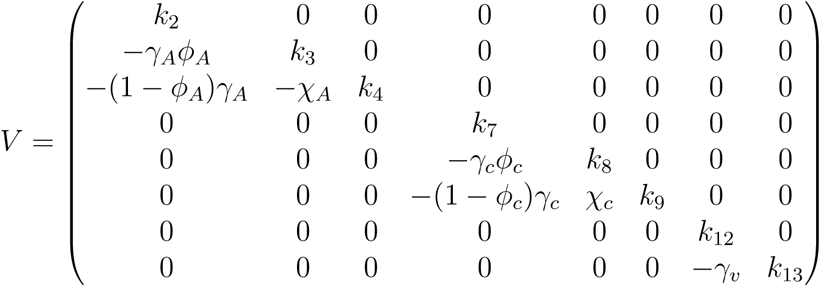

where,

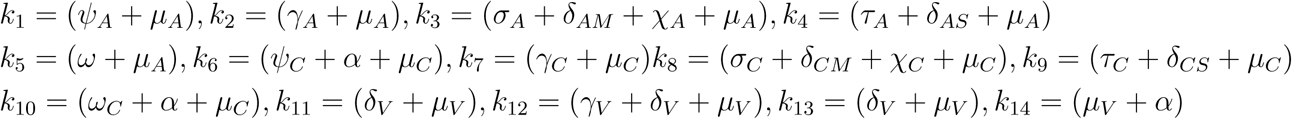

It follows that the effective reproduction number of model (1) denoted by *ℛ*_*E*_ is given by

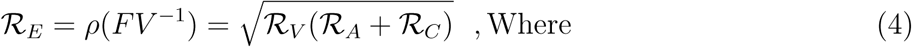

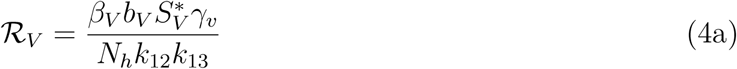

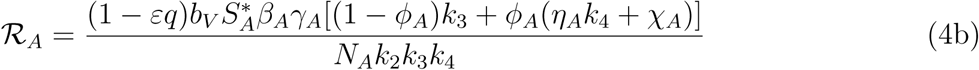

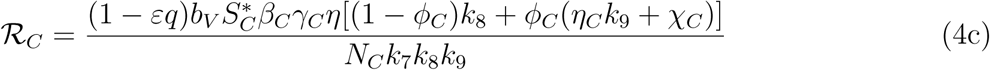

The result below follows from Theorem (2) in [12].

#### Lemma 3.

*The DFE ξ*_0_ *of model (1) is locally asymptotically stable (LAS) if ℛ*_*E*_ < 1, *and unstable if ℛ*_*E*_ > 1

The threshold quantity *ℛ*_*E*_, is the average number of malaria cases generated by a typically infected individual introduced into a completely susceptible population. The expression *ℛ*_*C*_ is the number of secondary infections in children introduced by one infectious mosquito, while the expression *ℛ*_*A*_ is the number of secondary infections in adult introduced by one infectious mosquito. The biological significance of lemma 3 is that malaria can be adequately controlled in a community with children and adults if the quantity *ℛ*_*E*_ can be reduced to a value less than unity (*ℛ*_*E*_ < 1)

### 3.2 Analysis of the Control Reproduction Number

The threshold quantity, will be used to determine the effect of the control parameters *ℛ*_*E*_ on the eradication of malaria in the population. For the sake of mathematical tractability in the analysis of our malaria transmission model, we shall work with the square of the reproduction number since our conclusion will not be altered if the actual expression of the reproduction number is used [13, 14]. From (4), we have

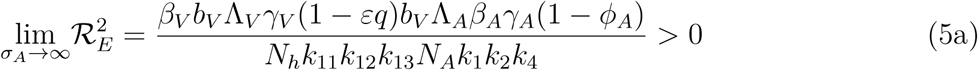

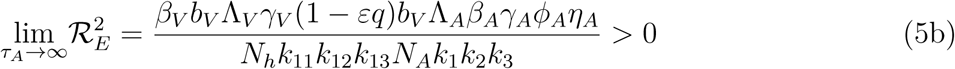

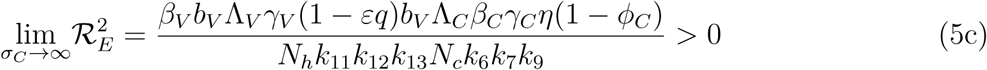

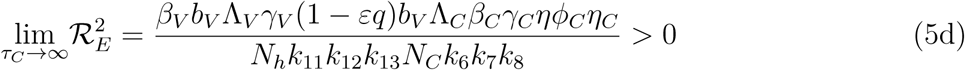

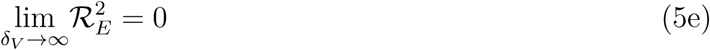

It therefore follows that control programme that results in high treatment rates and ITN compliance (*σ*_*A*_, *τ*_*A*_, *σ*_*c*_, *τ*_*c*_, *δ*_*v*_ → ∞) can lead to effective malaria control if these result in the respective right-hand sides of (5a-5d) being less than unity. From (5e), a near total eradication of malaria is achievable. This implies that focusing on reducing mosquito population through high rate of application of insecticide will serve as an effective malaria control strategy. Thus, differentiating the square of the reproduction number, 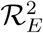, given in (4) partially with respect to the parameters (*σ*_*A*_, *τ*_*A*_, *σ*_*C*_, *τ*_*C*_), further reveals the effect of these parameters on malaria control in the community. Thus,

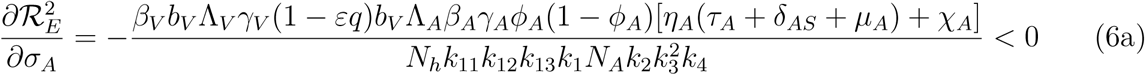

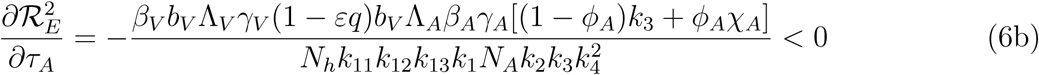

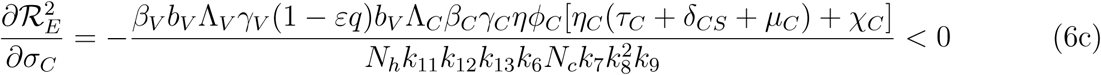

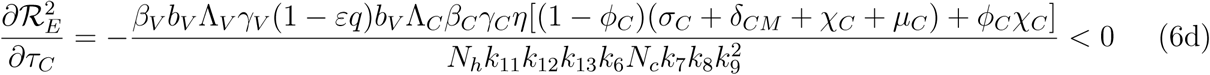

### 3.3 Assessing the Impact of the Mild Classes

Differentiating the square of the reproduction number, 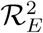, given in (4) partially with respect to the parameters (*η*_*A*_, *η*_*c*_) [13, 15] gives

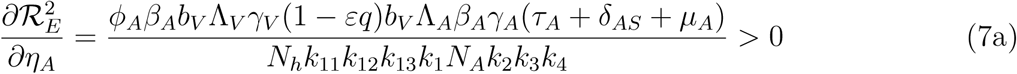

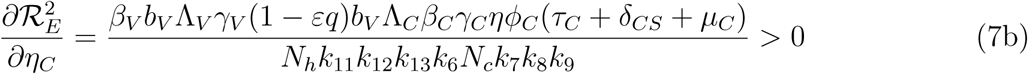

This means that the square of the effective reproduction number, 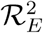, is an increasing function of the parameters (*η*_*A*_, *η*_*c*_). Thus, the disease burden of malaria in the community will increase as the infectivity of mildly infected children and adults increases. However, taking the limit of ^2^ as (*η*_*A*_, *η*_*c*_ → 1) implies that the infectivity of mildly infected children and adults is the same as that of the severely infected adults and children. Thus,

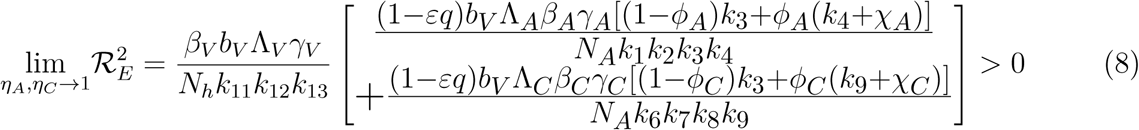

This implies that, as the infectivity of the mildly infected children and adults increases, the disease burden increases thereby increasing the number of malaria infected individuals in the community.

### 3.4 Existence of Endemic Equilibrium Point (EEP) of the Model

#### Lemma 4.

*The model (1) has a unique endemic(positive) equilibrium for the special case when ω* = *ω*_*C*_ = *α* = 0 *whenever ℛ*_*E*_ > 1

See **Appendix** for the proof of **Lemma 4**.

### 3.5 Bifurcation Analysis of the Model

It is important to explore the existence of bifurcation as this will go a long way in determining the parameter that will hinder the possibility of eradicating malaria transmission if the reproduction number is less than one. The centre manifold theorem is used here to investigate possibility of the existence of backward bifurcation as described [14, 16, 17].

#### Lemma 5.

*The transformed model (2) will undergo a backward bifurcation if the bifurcation coefficient a is positive*

See **Appendix** for the proof of **Lemma 5**.

### 3.6 Global Asymptotic Stability of DFE

#### Lemma 6.

*The DFE of the model (1) is globally asymptotically stable (GAS) in D whenever ℛ*_*E*_ ≤ 1

See **Appendix** for the proof of **Lemma 6**.

### 3.7 Global Asymptotic Stability of EEP

#### Lemma 7.

*The EEP of the model (1) is globally asymptotically stable (GAS) in D whenever ℛ*_*E*_ > 1

See **Appendix** for the proof of **Lemma 7**.

## 4 Analysis of Control Strategies

To effectively study the behavioral pattern of malaria transmission, some important parameters of the model are explored here to quantify the effectiveness of malaria intervention strategies. We now carry out numerical analysis to investigate the relative importance of the parameters of our model in disease transmission and how best to tackle malaria outbreak and reduce malaria mortality. In particular, we target intervention strategies using parameters such as mosquito recruitment rate (Λ_*V*_), mosquito death rate (*δ*_*V*_), mosquito biting rates (*b*_*V*_), rate of ITN compliance (*q*), vaccination rates (*ψ*_*A*_, *ψ*_*C*_) and treatment rates (*σ*_*A*_, *τ*_*A*_, *σ*_*C*_, *τ*_*C*_). Theoretical values are used to represent parameter values and initial conditions in our simulations since they are similar to comparable parameters for other mosquito-transmitted diseases [13, 14].

### 4.1 Mosquito-Reduction Strategy

The use of indoor residual spraying (IRS) and DDT reduce the average lifespan(*δ*_*V*_)of mosquitoes and mosquito recruitment rate/birth rate (Λ_*V*_)respectively. The following three levels of mosquito-reduction strategies are considered for simulation [13, 15]

**Table 4.1:** Description of parameter values

| Parameter | Values | Range | References |
| --- | --- | --- | --- |
| $\Lambda_A$ | 235 | 100 – 500 | Estimated |
| $\Lambda_C$ | 156 | 100 – 500 | Estimated |
| $\Lambda_V$ | 16667 | 7000 – 40000 | Estimated |
| $\mu_A$ | $\frac{1}{70 \times 365}$ | $\frac{1}{76 \times 365} - \frac{1}{68 \times 365}$ | [13] |
| $\mu_C$ | $\frac{1}{18.60 \times 365}$ | $\frac{1}{14.88 \times 365} - \frac{1}{22.32 \times 365}$ | [1] |
| $\mu_V$ | $\frac{1}{21}$ | $\frac{1}{42} - \frac{1}{8}$ | [18], [19] |
| $\beta_A, \beta_C$ | 0.017, 0.14 | 0.01 – 0.27 | [8] |
| $\beta_V$ | 0.356 | 0.072 – 0.64 | [8] |
| $\gamma_A, \gamma_C$ | 0.134, 0.122 | 0.067 – 0.20 | [8] |
| $\phi_A, \phi_C, \psi_A, \psi_C$ | 0.5 | 0.1 – 1.0 | Assumed |
| $\delta_{AM}, \delta_{AS}$ | 0.00021, 0.0046 | 0.0 – 0.00041 | [8] |
| $\delta_{CM}, \delta_{CS}$ | 0.0003454, 0.00671 | 0.000135 – 0.00817 | [10] |
| $\delta_V$ | 0.5 | 0.25 – 0.75 | [13] |
| $\sigma_A, \tau_A$ | 0.0099, 0.014 | 0.0014 – 0.017 | [8] |
| $\sigma_C, \tau_C$ | 0.0082, 0.011 | 0.0014 – 0.017 | [8] |
| $\theta$ | 0.4 | 0.1 – 1.0 | Assumed |
| $\omega$ | 0.0243 | 0.0025 – 0.067 | [10] |
| $\alpha$ | $\frac{1}{16 \times 365}$ | $\frac{1}{18 \times 365} - \frac{1}{15 \times 365}$ | [13] |
| $\omega_C$ | 0.046 | 0.0025 – 0.067 | Estimated |
| $\gamma_V$ | 0.18 | 0.029 – 0.33 | [8] |
| $b_V$ | 0.5 | 0.1 – 1.0 | [13] |
| $\varepsilon; 0 \leq \varepsilon \leq 1$ | 0.5 | 0.2 – 1.0 | [9] |
| $q$ | 0.53 | 0.1325 – 0.6625 | [9] |
| $\eta$ | 0.5 | 0.1 – 1.0 | Assumed |
| $\eta_A, \eta_C$ | 0.5 | 0.1 – 1.0 | Assumed |
| $\chi_A, \chi_C$ | 0.091, 0.11 | 0.067 – 0.20 | [8] |

1. Low mosquito-reduction strategy Λ_*V*_ = 33334*/day*; *δ*_*V*_ = 0.25 */day*
2. Moderate mosquito-reduction strategy Λ_*V*_ = 16667*/day*; *δ*_*V*_ = 0.5 */day*
3. High mosquito-reduction strategy Λ_*V*_ = 8334*/day*; *δ*_*V*_ = 0.75 */day*

Fig (1a) and (1c) indicate a decreasing pattern in both adult and children population with mild malaria while varying the recruitment rate of mosquitoes whereas Fig (1b) and (1d) show that adult and children population with severe malaria increase to a maximum value before they start decreasing gradually while varying mosquito recruitment rate.

**Figure 1:**
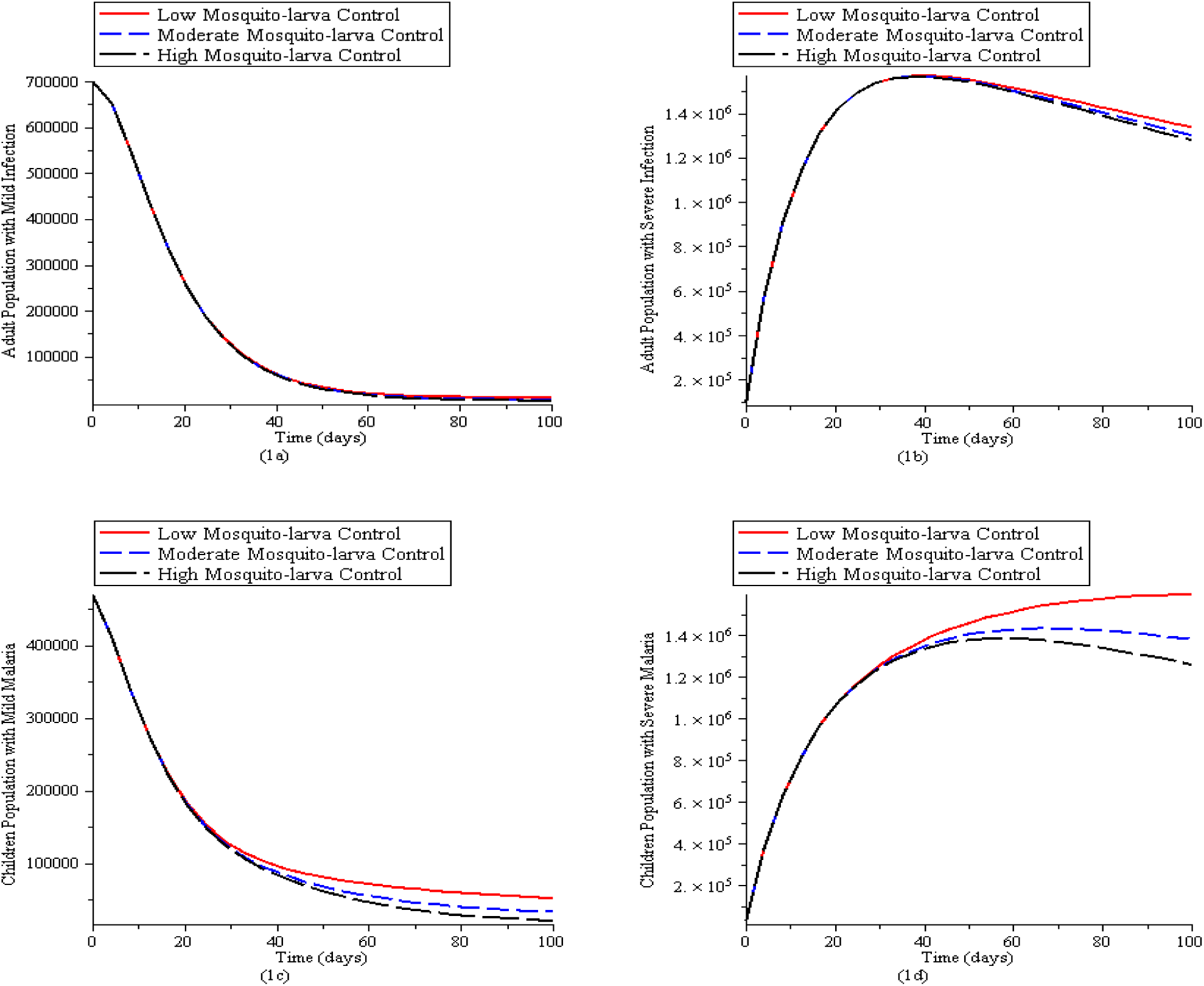
Simulation of the Malaria model (1) for various control levels of the mosquito reduction strategy. (1a). New cases of adult population with mild malaria. (1b). New cases of adult population with severe malaria. (1c). New cases of children population with mild malaria. (1d) New cases of children population with severe malaria. The parameter values used are in Table2.1 with *q* = 0, *ψ*_*A*_ = 0, *ψ*_*C*_ = 0, *δ*_*V*_ = 0, *σ*_*A*_ = 0, *σ*_*C*_ = 0, *τ*_*C*_ = 0, *τ*_*A*_ = 0,

Fig (2a) and (2c) indicate a decreasing pattern in both adult and children population with mild malaria irrespective of the level of mosquitoes insecticide spray applied whereas Fig (2b) and (2d) show that high effectiveness mosquito reduction strategy lead to a considerable reduction in the number of severe malaria cases in adult and children population compared to the moderate-effectiveness level [13, 15].

**Figure 2:**
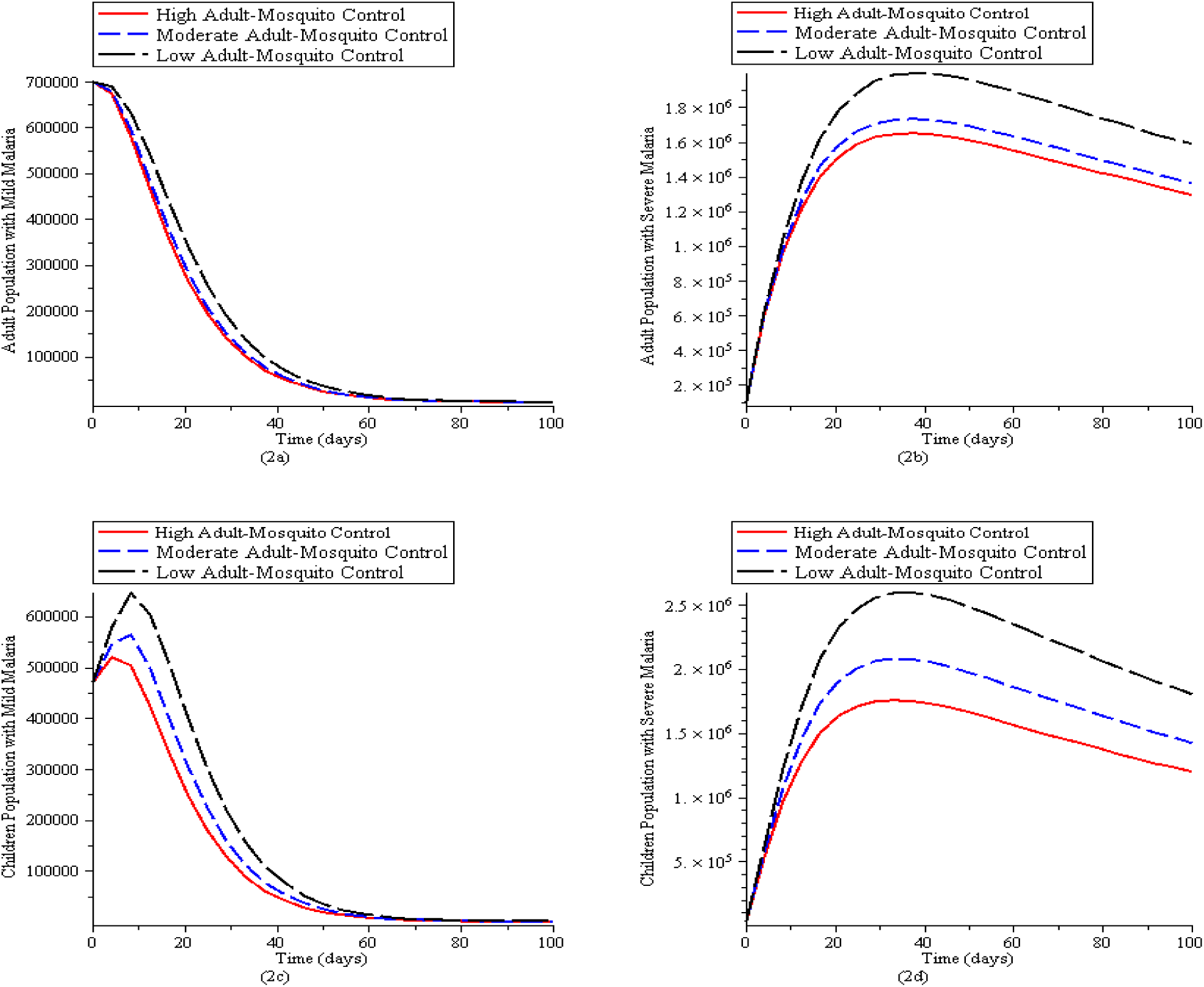
Simulation of the Malaria model (1) for various control levels of the mosquito reduction strategy. (2a). New cases of adult population with mild malaria. (2b). New cases of adult population with severe malaria. (2c). New cases of children population with mild malaria. (2d) New cases of children population with severe malaria. The parameter values used are in Table 2.1 with *q* = 0, *ψ*_*A*_ = 0, *ψ*_*C*_ = 0, *σ*_*A*_ = 0, *σ*_*C*_ = 0, *τ*_*C*_ = 0, *τ*_*A*_ = 0,

### 4.2 Personal Protection Strategy

Personal protection involving the use of ITN (*q*)reduces the exposure rate of humans to mosquitoes which in turn reduces biting rates (*b*_*V*_) and the transmission of parasites between humans and (*b*_*V*_) mosquitoes. We consider the following three levels [13];

1. Low personal protection strategy *q* = 0.133*/day*; *b*_*V*_ = 0.75 */day*
2. Moderate personal protection strategy *q* = 0.265*/day*; *b*_*V*_ = 0.50 */day*
3. High personal protection strategy *q* = 0.53*/day*; *b*_*V*_ = 0.25 */day*

In Fig (3a) and (3d),mild malaria cases in adult reduces faster within a shorter period as compared to mild malaria cases in children whereas Fig (3b) and (3c) show that with high personal protection strategy (use of insecticide treated nets) the number of severe malaria cases in adult and children population increase considerably to a level before decreasing gradually.

**Figure 3:**
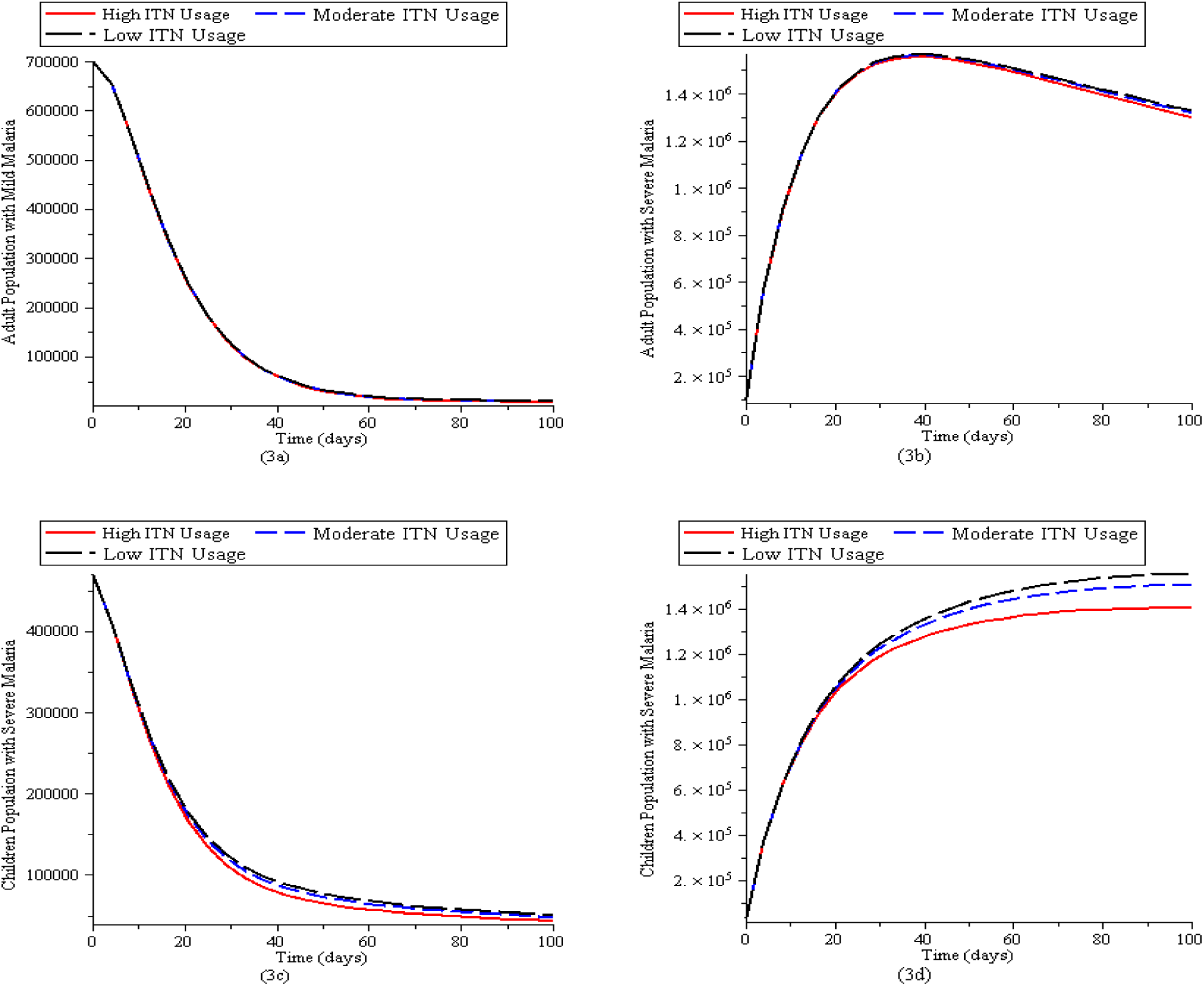
Simulation of the Malaria model (1) for various control levels of the mosquito reduction strategy. (3a). New cases of adult population with mild malaria. (3b). New cases of adult population with severe malaria. (3c). New cases of children population with mild malaria. (3d) New cases of children population with severe malaria. The parameter values used are in Table 2.1 with *ψ*_*A*_ = 0, *ψ*_*C*_ = 0, *δ*_*V*_ = 0, *σ*_*A*_ = 0, *σ*_*C*_ = 0, *τ*_*C*_ = 0, *τ*_*A*_ = 0,

In Fig (4a) and (4c), mild malaria cases in adult reduces faster within a shorter period as compared to mild malaria cases in children except when the biting rate is low in children population whereas Fig (4b) and (4d) show that when there is low personal protection strategy (high biting rate)the children population with severe malaria will increase more compared to the moderate-effectiveness level and high-effectiveness level.

**Figure 4:**
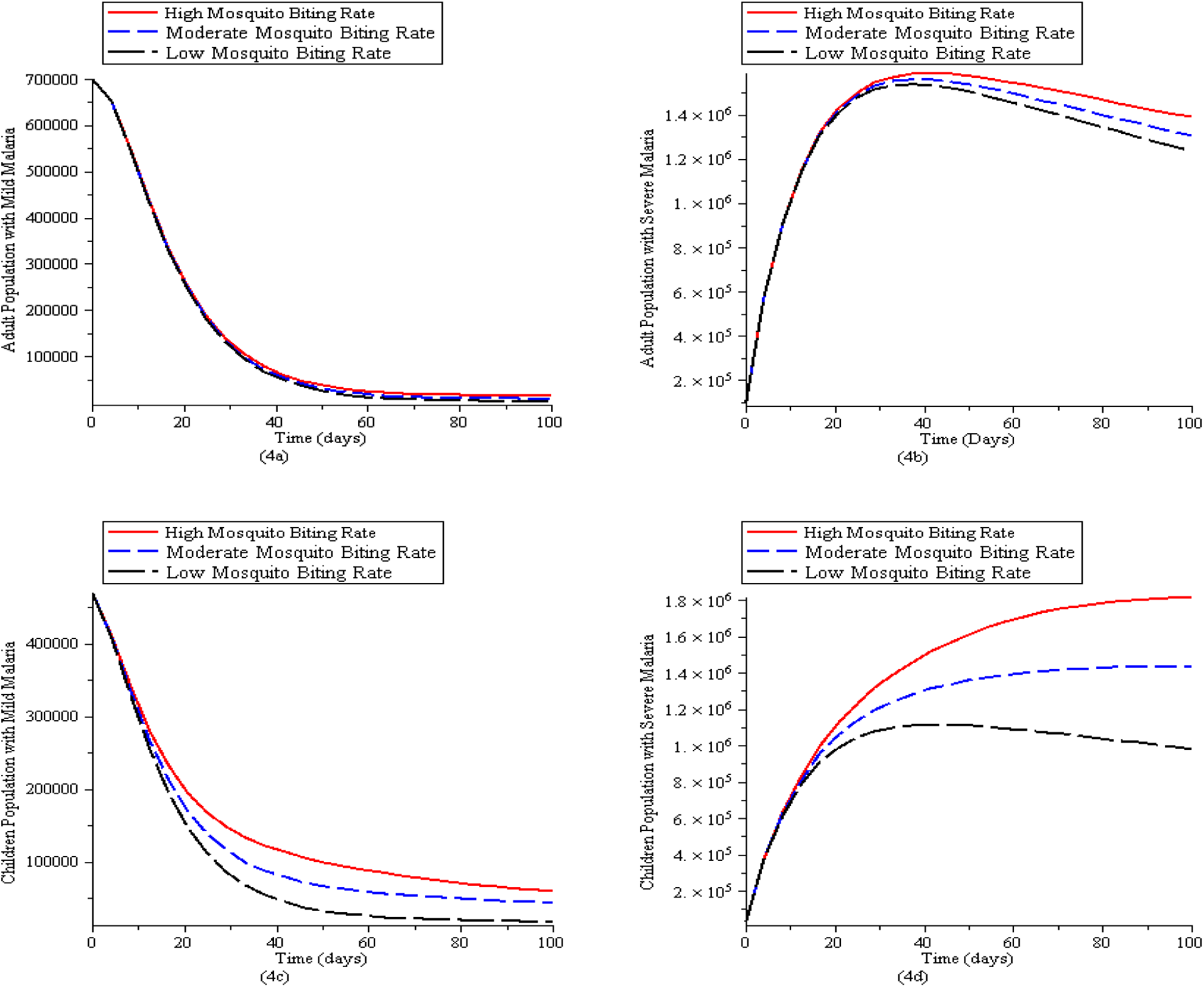
Simulation of the Malaria model 1 for various control levels of the mosquito reduction strategy. (4a). New cases of adult population with mild malaria. (4b). New cases of adult population with severe malaria. (4c). New cases of children population with mild malaria. (4d) New cases of children population with severe malaria. The parameter values used are in Table2.1 with *q* = 0, *ψ*_*A*_ = 0, *ψ*_*C*_ = 0, *δ*_*V*_ = 0, *σ*_*A*_ = 0, *σ*_*C*_ = 0, *τ*_*C*_ = 0, *τ*_*A*_ = 0

### 4.3 Vaccination Strategy

1. Low vaccination strategy *ψ*_*A*_ = 0.25*/day*; *ψ*_*C*_ = 0.25*/day*
2. Moderate vaccination strategy *ψ*_*A*_ = 0.5*/day*; *ψ*_*C*_ = 0.5*/day*
3. High vaccination strategy *ψ*_*A*_ = 0.75*/day*; *ψ*_*C*_ = 0.75*/day*

In Fig (5a) and (5b), susceptible adult and children population reduces faster due to vaccination whereas Fig (5c) and (5d) show that vaccination increases the immune adult and children population.

**Figure 5:**
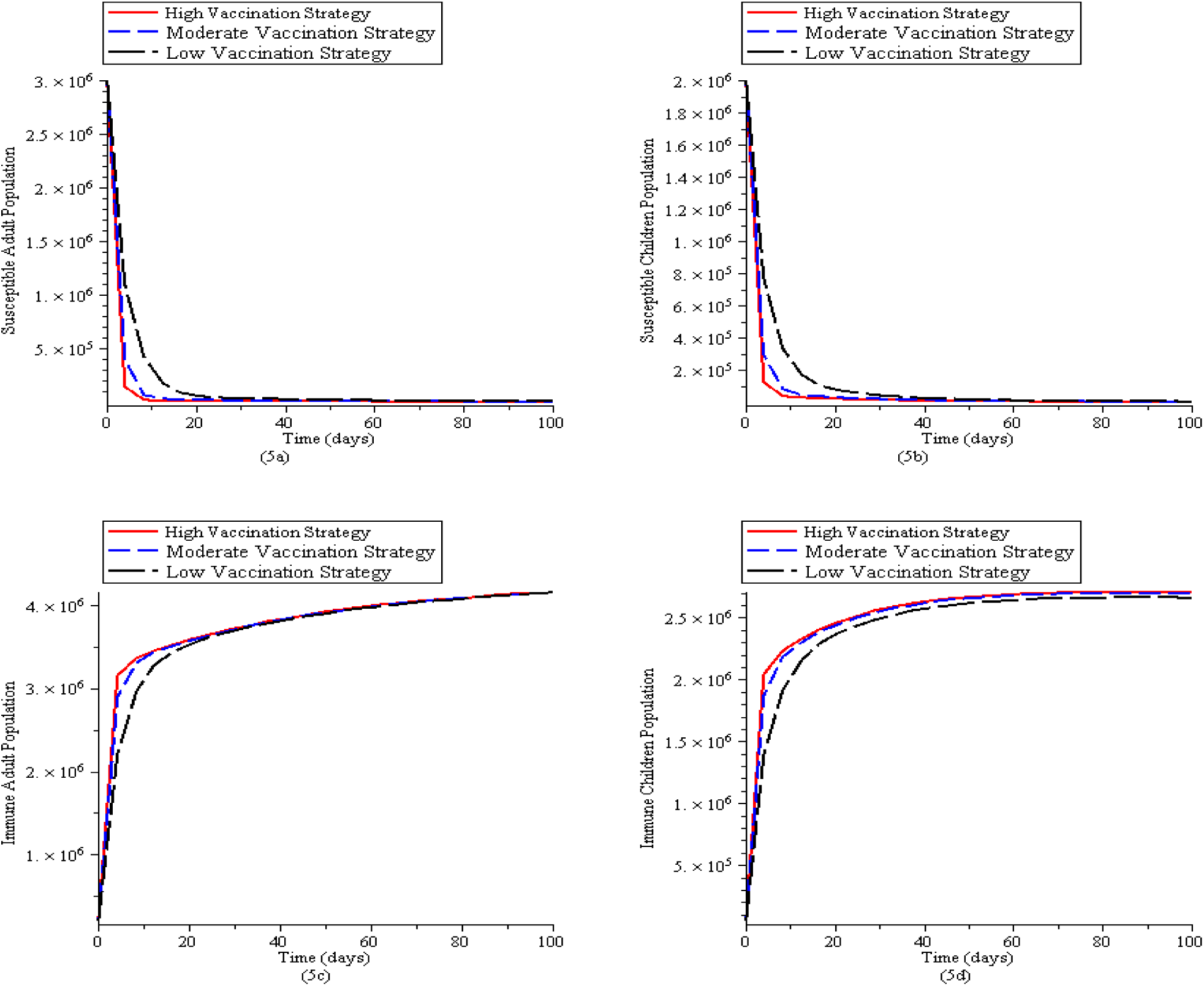
Simulation of the Malaria model (1) for various control levels of the mosquito reduction strategy. (5a). New cases of susceptible adult population. (5b). New cases of susceptible children population. (5c). New cases of immune adult population (5d) New cases of immune children population. The parameter values used are in Table 2.1 with *q* = 0, *δ*_*V*_ = 0, *σ*_*A*_ = 0, *σ*_*C*_ = 0, *τ*_*C*_ = 0, *τ*_*A*_ = 0

### 4.4 Combined Strategy

This strategy combines the mosquito reduction, personal protection and vaccination strategy under the following three control levels;

1. Low Combined Control Λ_*V*_ = 33334*/day*; *δ*_*V*_ = 0.25*/day*; *q* = 0.133*/day*; *b*_*V*_ = 0.75*/day*; *ψ*_*A*_ = 0.25*/day*; *ψ*_*C*_ = 0.25*/day*
2. Moderate Combined Control Λ_*V*_ = 16667*/day*; *δ*_*V*_ = 0.5*/day*; *q* = 0.265*/day*; *b*_*V*_ = 0.50*/day*; *ψ*_*A*_ = 0.5*/day*; *ψ*_*C*_ = 0.5*/day*
3. High Combined Control Λ_*V*_ = 8334*/day*; *δ*_*V*_ = 0.75*/day*; *q* = 0.53*/day*; *b*_*V*_ = 0.25*/day*; *ψ*_*A*_ = 0.75*/day*; *ψ*_*C*_ = 0.75*/day*

In Fig (6a), (6b), (6c) and (6d), the presence of all the control strategies is more effective in preventing mild malaria cases in adult and children as compared to severe malaria cases in adult and children.

**Figure 6:**
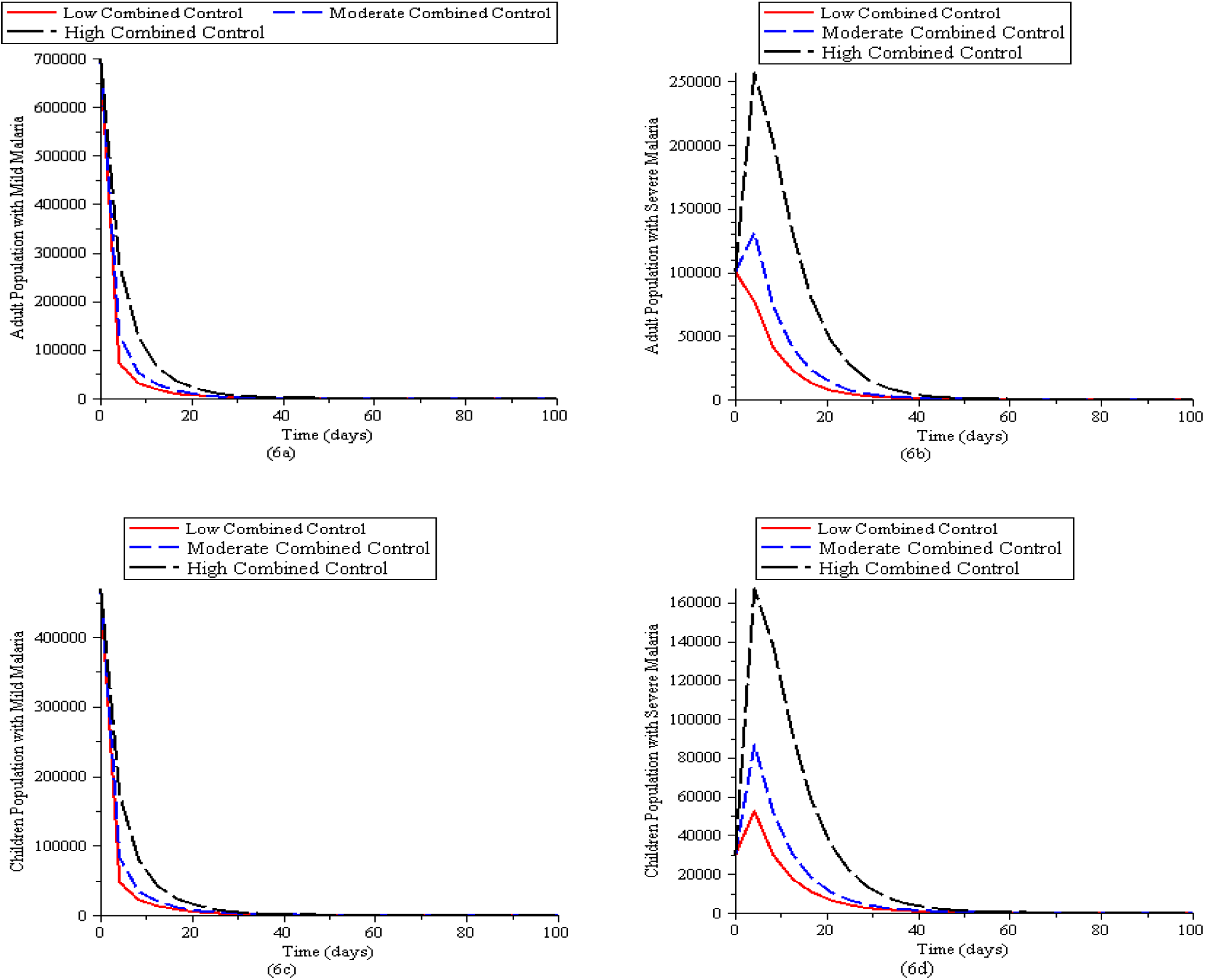
Simulation of the Malaria model (1) for various control levels of the mosquito reduction strategy. (6a). New cases of adult population with mild malaria. (6b). New cases of adult population with severe malaria. (6c). New cases of children population with mild malaria. (6d) New cases of children population with severe malaria. The parameter values used are in Table 2.1

## 5 Discussion and Conclusion

For decades now, deliberate policies have been formulated to prevent and reduce the transmission of malaria with some degree of success recorded in some developed parts of the world. In this paper, we formulated and analysed a mathematical model of malaria in a heterogeneous population incorporating mosquito reduction strategy, personal protection strategy and vaccination as control strategies. These control strategies are designed to reduce the contact rates between humans and mosquitoes. Some epidemiological findings of this study are summarized below:

1. The Disease Free Equilibrium point (DFE) of the malaria model (1) is locally asymptotically stable (LAS) if *ℛ*_*E*_ < 1(unstable if *ℛ*_*E*_ > 1) and globally asymptotically stable (GAS) in D whenever *ℛ*_*E*_ ≤ 1
2. The model (1) has a unique endemic(positive) equilibrium for the special case when *ω* = *ω*_*C*_ = *α* = 0 whenever*R*_*E*_>1
3. The endemic equilibrium point of model (1) is globally asymptotically stable (GAS) if *ℛ*_*E*_>1
4. The model (1) will undergo a backward bifurcation whenever a stable disease free equilibrium point coexists with a stable endemic equilibrium point when the associated reproduction number is less than unity. However, the model (1) does not undergo the phenomenon of backward bifurcation if *ω*(1 − *θ*) = *ω*_*C*_ = *η*_*A*_ = *α* = 0 Hence, this study shows that the loss of acquired immunity of recovered adults (*ω*(1 − *θ*)), loss of acquired immunity of recovered children (*ω*_*C*_) and the loss of likelihood (*η*_*A*_) of adults getting mild infection are the causes of backward bifurcation in the malaria transmission model. However, the presence of *α* in the bifurcation coefficient *a* indicates that imbalance in growth and maturation from childhood to adulthood can equally cause backward bifurcation. Hence the model does not undergo the phenomenon of backward bifurcation so long as the recovered children and adults do not lose their acquired immunity, if the infection of mildly infected adult is not high enough to infect susceptible mosquitoes and if there is proper balance in the factors affecting growth and maturation. These growth factors include heredity, environment, exercise & health, hormones, nutrition, familial influence, geographical influence, socio-economic status, learning and reinforcement.
5. From analysis of control reproduction number, focusing on reducing mosquito population through high rate of application of insecticide will serve as an effective malaria control strategy.
6. From numerical simulation the following results were obtained
  i. mild malaria cases are easier to control in both adult and children as compared to the severe cases.
  ii. high mosquito biting rate causes more harm in children population than in adult population.
  iii. the presence of all the control strategies is more effective in preventing mild malaria cases in adult and children as compared to severe malaria cases in adult and children.
7. Therefore, more effort should be put in place to prevent severe malaria cases in the population

## Data Availability

Details on data source are included in the manuscripts. Sources are properly referenced.

## Appendix

***Proof:***(Proof of Lemma 4)

The existence of endemic equilibrium of model (1) for the special case when *ω* = *ω*_*C*_ = *α* = 0 is established as follows.

Let the EEP of the model be

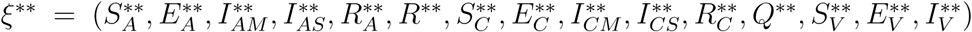.. The equations in (1) with *ω* = *ω*_*C*_ = *α* = 0 is solved in terms of the forces of infection at the steady state to give

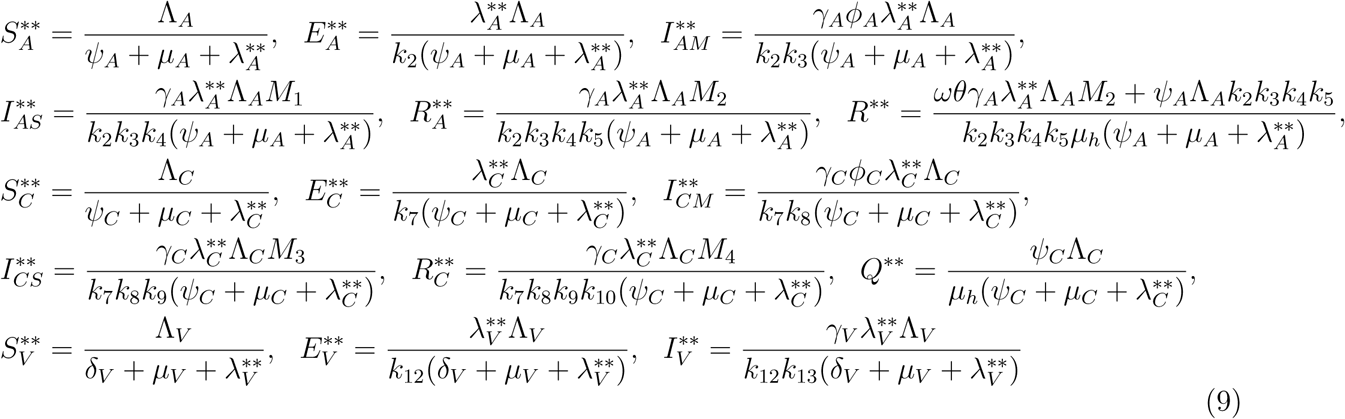

Where

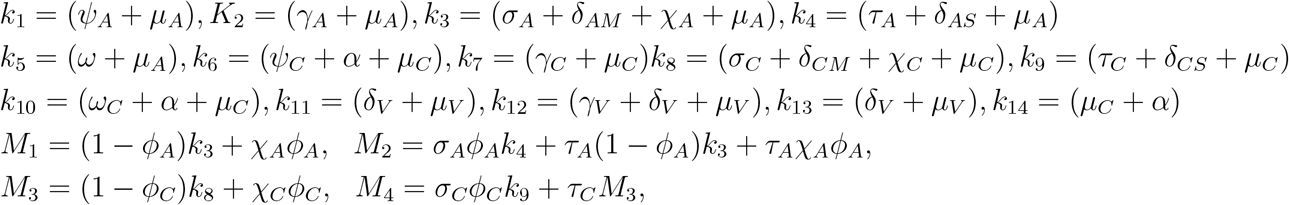

Substituting the values of 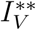 into 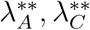 gives

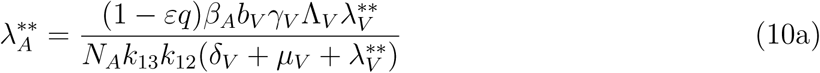

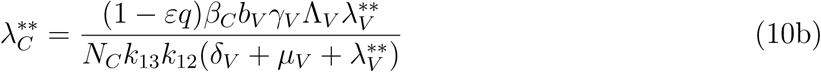

Substituting the values of 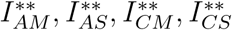 into 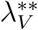 gives

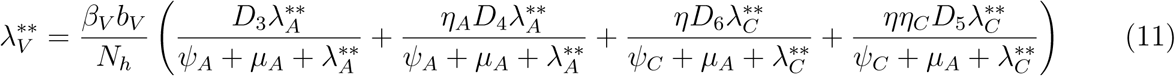

Where

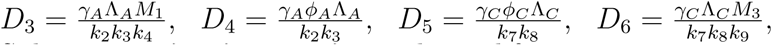

Substituting 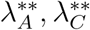 into 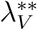 and simplifying gives

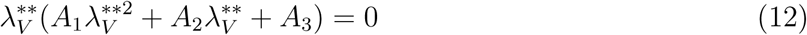

Which implies either 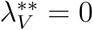 or Where

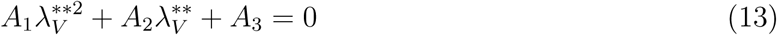

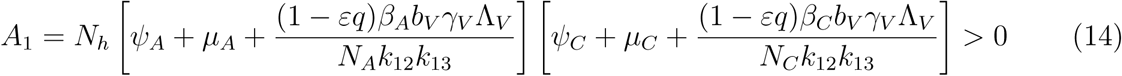

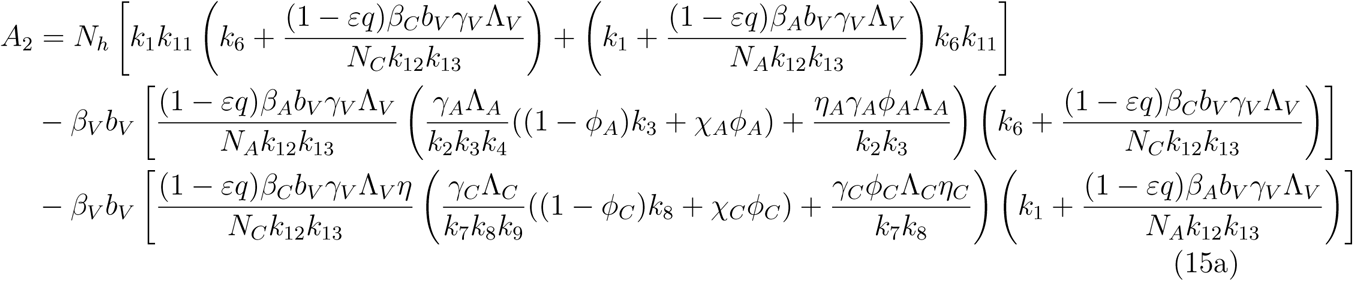

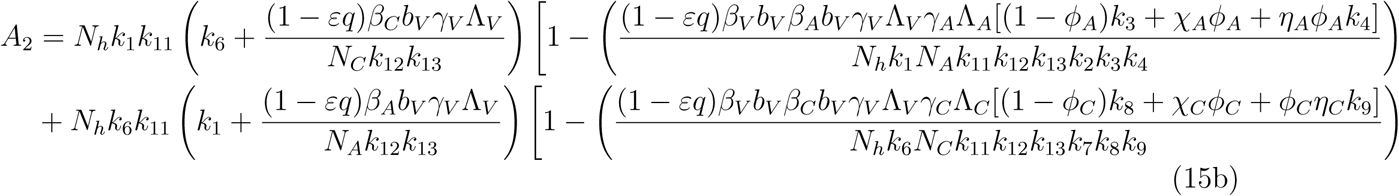

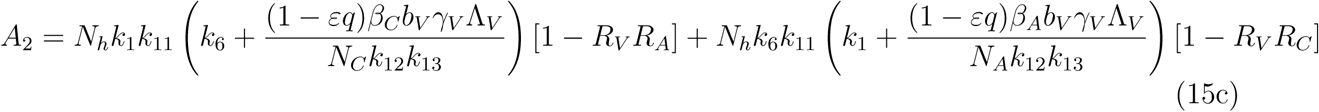

*A*_2_ < 0 if ℛ_*V*_ℛ_*A*_ > 1 and ℛ_*V*_ℛ_*C*_, *A*_2_ > 0 if ℛ_*V*_ℛ_*A*_ < 1 and ℛ_*V*_ℛ_*C*_ < 1

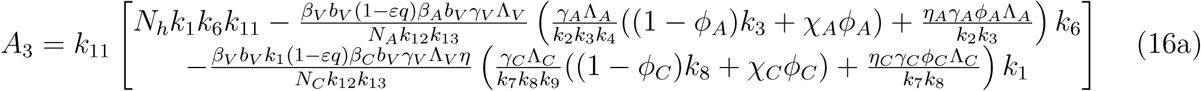

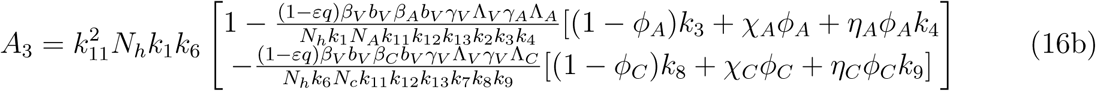

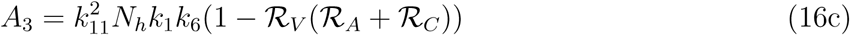

*A*_*3*_ > *0 if ℛv(ℛ*_*A*_ *+ ℛ*_*C*_) < *1*

*A*_*3*_ < *0 if ℛv(ℛ*_*A*_ *+ ℛ*_*C*_) < *1*

***Proof:***(Proof of Lemma 5)

For model (1), let

(*x*_1_ = *S*_*A*_, *x*_2_ = *E*_*A*_, *x*_3_ = *I*_*AM*_, *x*_4_ = *I*_*AS*_, *x*_5_ = *R*_*A*_, *x*_6_ = *R, x*_7_ = *S*_*C*_, *x*_8_ = *E*_*C*_, *x*_9_ = *I*_*CM*_, *x*_10_ = *I*_*CS*_, *x*_11_ = *R*_*C*_, *x*_12_ = *Q, x*_13_ = *S*_*V*_, *x*_14_ = *E*_*V*_, *x*_15_ = *I*_*V*_).

Thus, the model can be written as

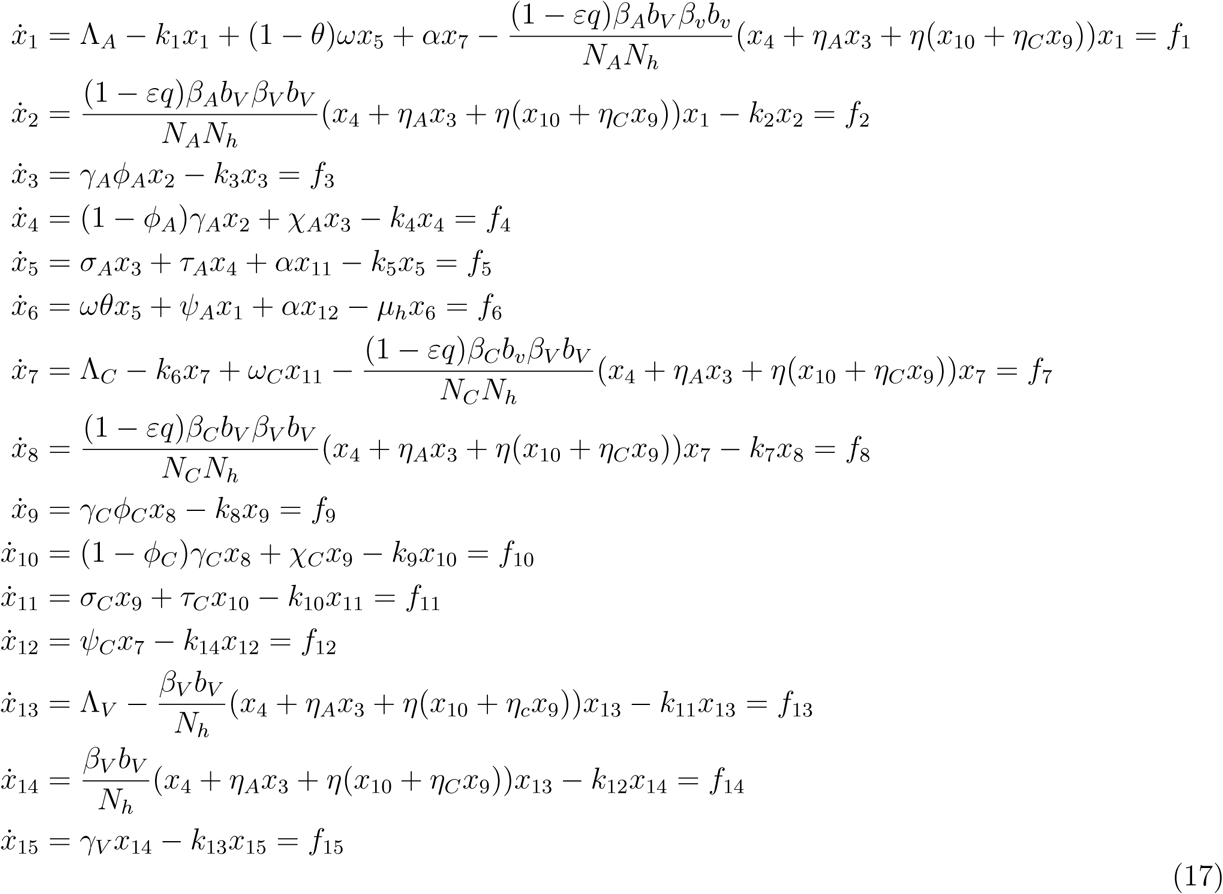

Consider the case when 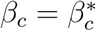 is chosen as the bifurcation parameter. Solving for 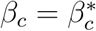 from *ℛ*_*E*_ = 1 gives

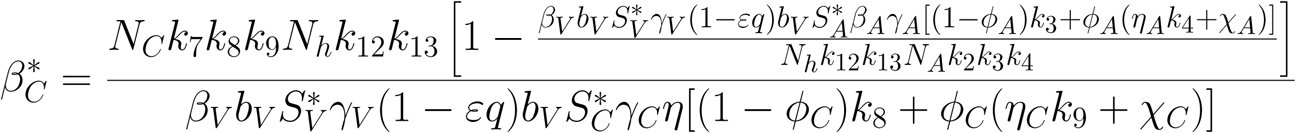

The Jacobian *J* (*ξ*^*\**^) of the transformed system (17) evaluated at the disease-free equilibrium with 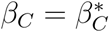 gives

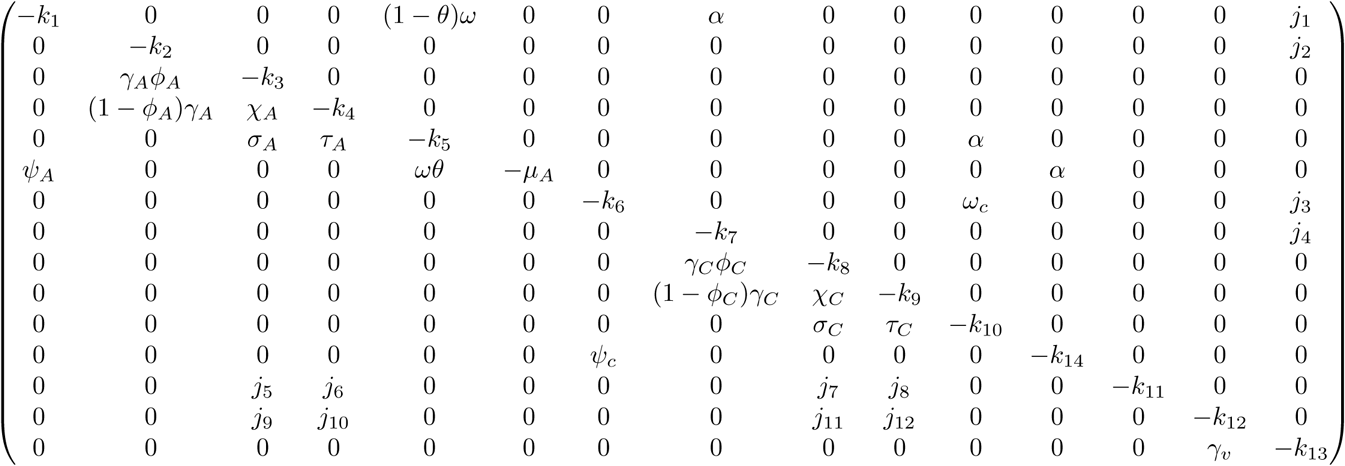

where,

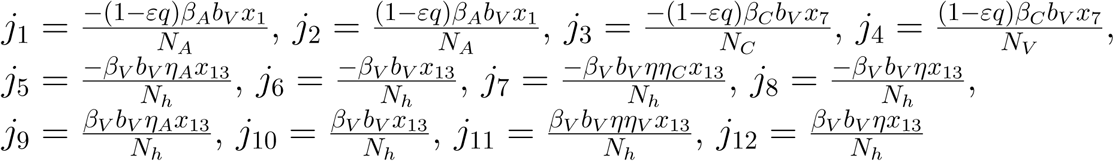

The right eigen vector of *J* (*ξ*^*\**^) is given by *w* = (*w*_1_, *w*_2_, …*w*_15_)^*T*^, where

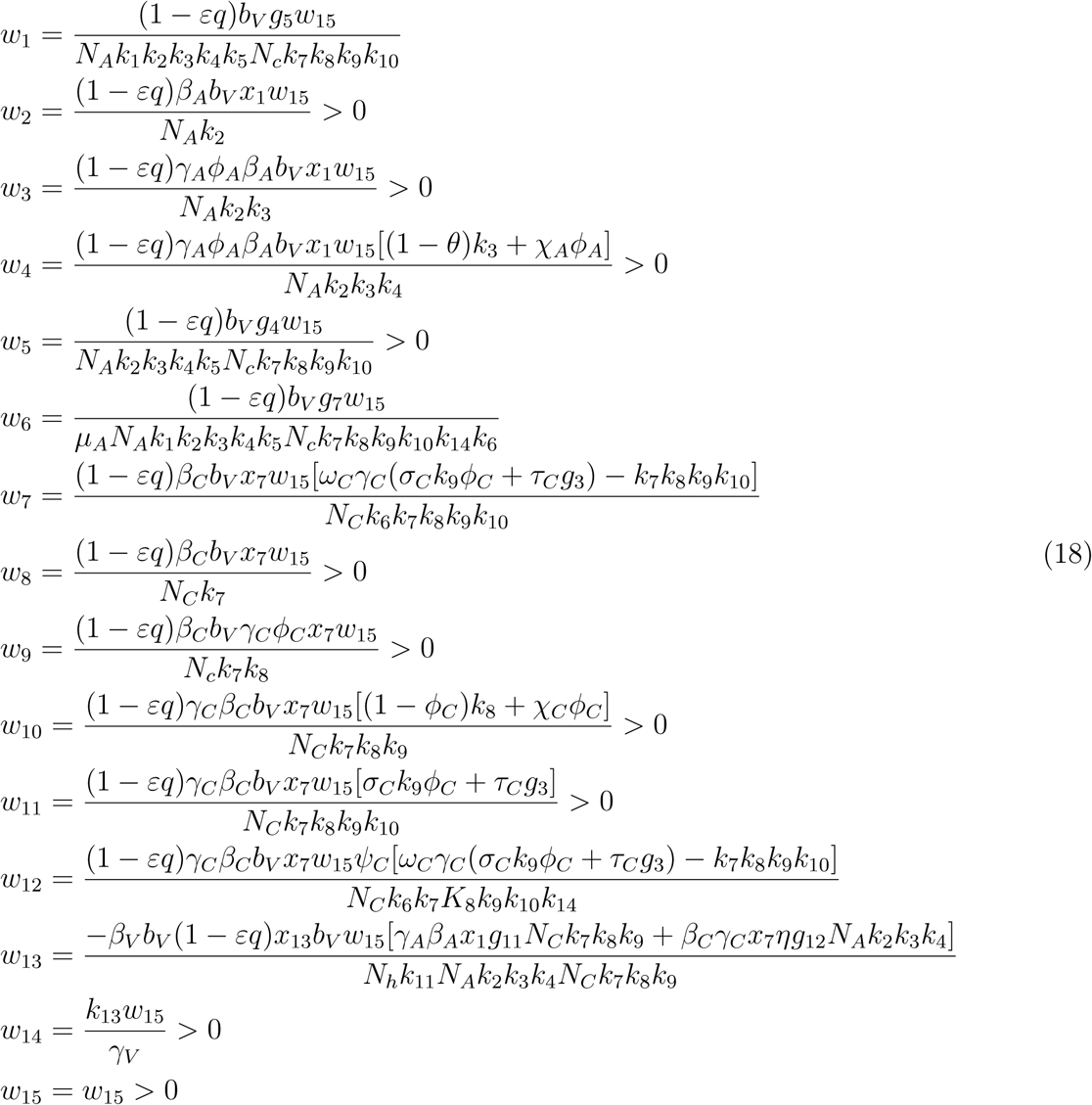

Where

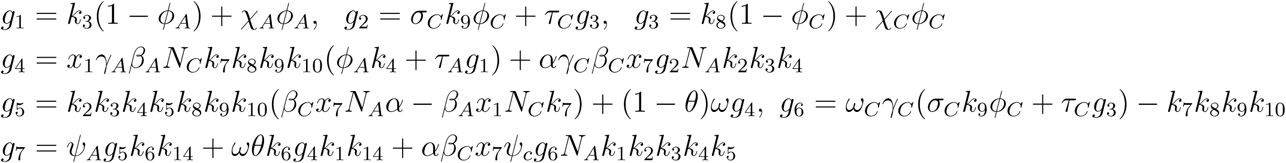

Similarly, 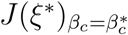 has a left eigenvector *v* = (*v*_1_, *v*_2_, …, *v*_15_), where

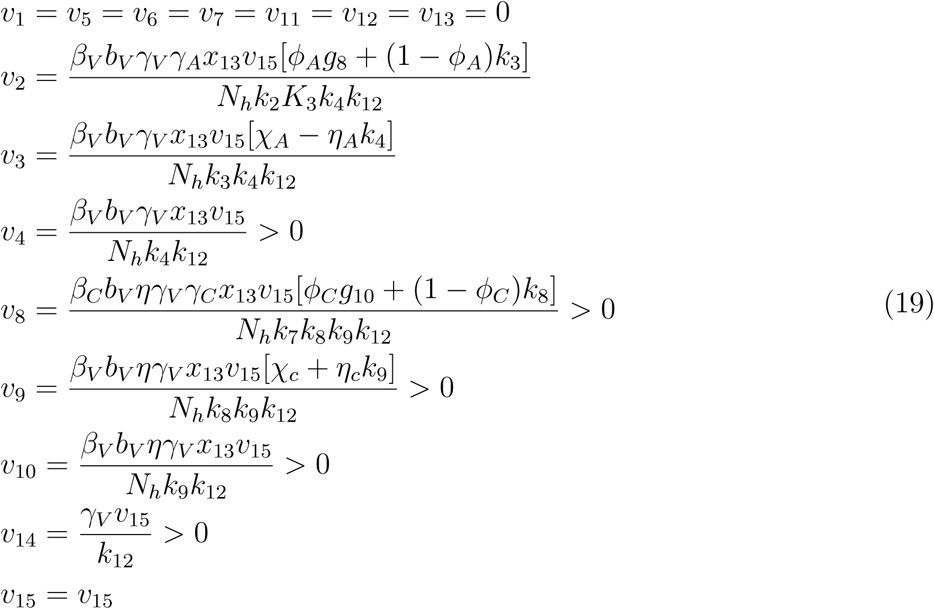

Where

*g*_8_ = *χ*_*A*_ *η*_*A*_*k*_4_, *g*_10_ = *χ*_*C*_ *η*_*C*_*k*_9_, *g*_11_ = *ϕ*_*A*_*η*_*A*_*k*_4_ + *k*_3_(1 − *ϕ*_*A*_) + *χ*_*A*_*ϕ*_*A*_, *g*_12_ = *ϕ*_*C*_*η*_*C*_*k*_9_ + *g*_3_

The associated non-zero partial derivatives of system (17) evaluated at the DFE gives the associated bifurcation coefficients *a* and *b* defined by

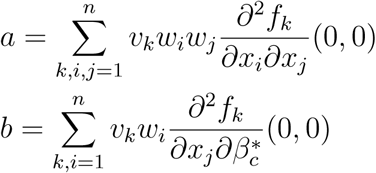

which gives

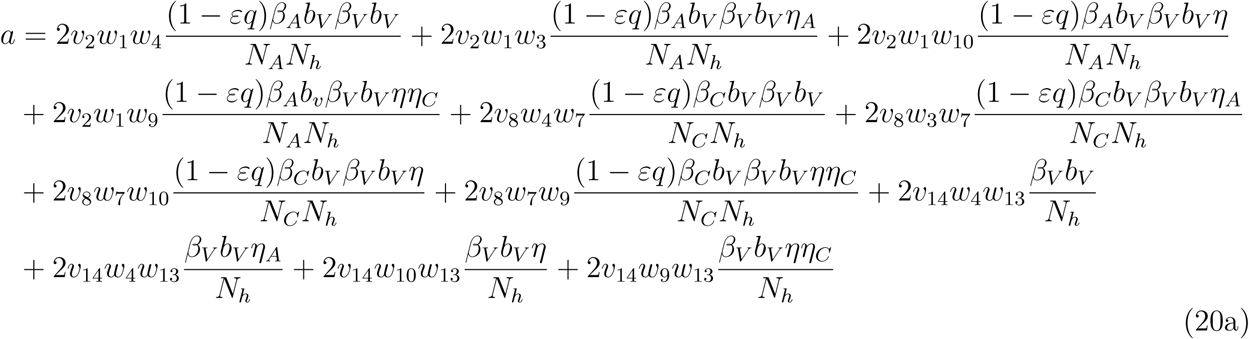

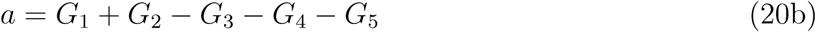

where

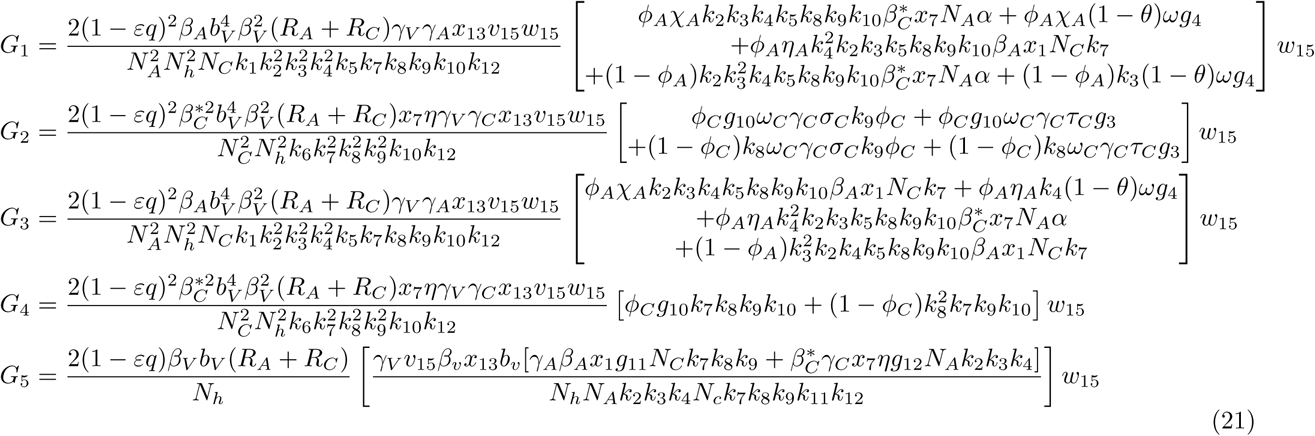

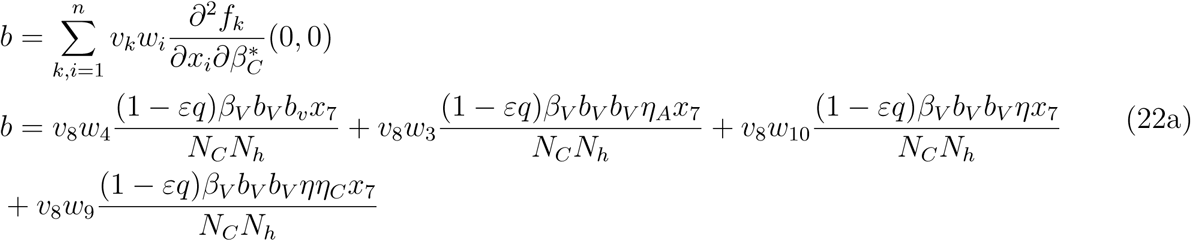

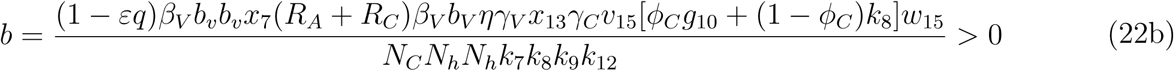

Since the bifurcation coefficient is positive. It follows from theorem 2 of ([16]) that the transformed model (17) will undergo a backward bifurcation if the bifurcation coefficient *a* is positive. The phenomenon of backward bifurcation examines the scenario where a stable DFE coexist with a stable EEP when the associated reproduction number is less than unity.

The epidemiological implication of the backward bifurcation of the model (1) is that the classical requirement of the reproduction number being less than unity becomes only a necessity, but not sufficient condition for malaria control. However, if we set *ω*(1 − *θ*) = *ω*_*C*_ = *η*_*A*_ = *α* = 0 in the expression for *a* in (20b), the bifurcation parameter becomes negative. Thus, it follows from the *a* Castillo Chavez theorem in [16], that model (1) does not undergo the phenomenon of backward bifurcation if *ω*(1 − *θ*) = *ω*_*C*_ = *η*_*A*_ = *α* = 0.

***Proof:***(Proof of Lemma 6)

To prove the global asymptotic stability of DFE we use a previously described approach in [13]. Let *X* = (*S*_*A*_, *R*_*A*_, *R, S*_*C*_, *R*_*C*_, *Q, S*_*V*_) and *Z* = (*E*_*A*_, *I*_*AM*_, *I*_*AS*_, *E*_*C*_, *I*_*CM*_, *I*_*CS*_, *E*_*V*_, *I*_*V*_) and writing the model equation (1) in the form

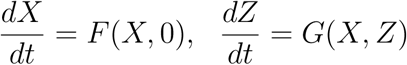, where *E*_*A*_, *I*_*AM*_ *= I*_*AS*_ *= E*_*C*_*= I*_*CM*_ *= I*_*CS*_ *= E*_*V*_ *= I*_*V*_ *=0* with *F* (*X*, 0) being the RHS of 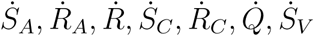 and *G*(*X, Z*) the RHS of 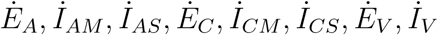

Next, consider the reduced system: 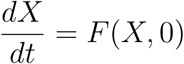 given as

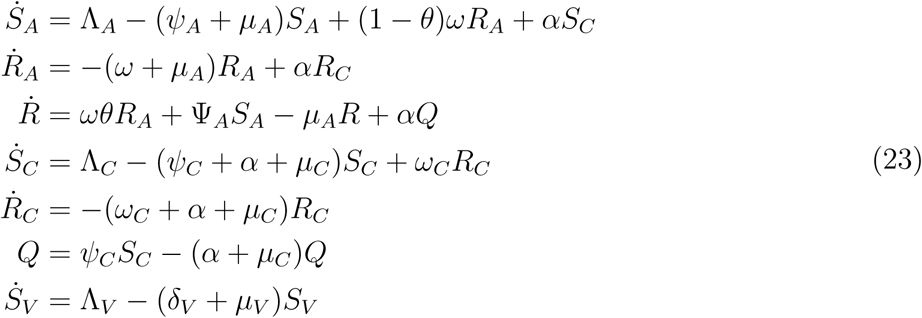

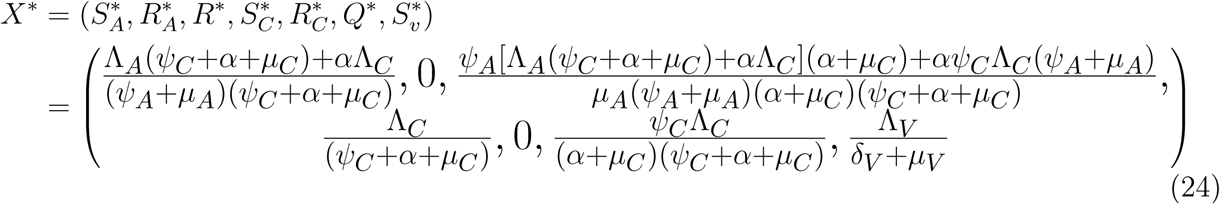

be an equilibrium of the reduced system (23), we now show that *X*^*\**^ is a globally stable equilibrium in *D* by solving equations (23) and taking limit as *t* → ∞

Solving for *S*_*A*_(*t*) gives

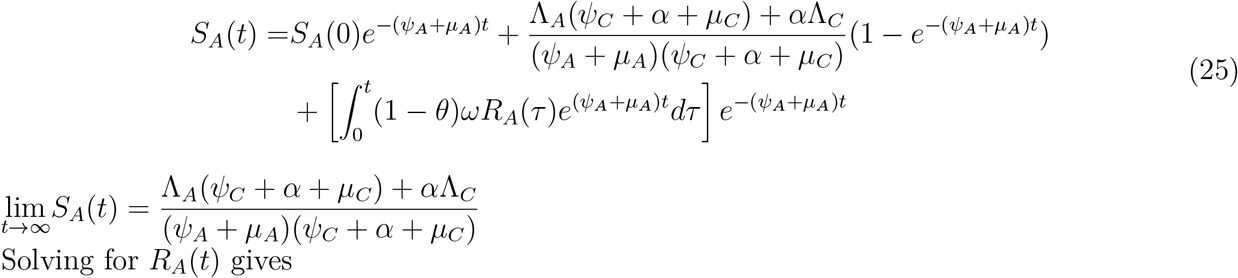

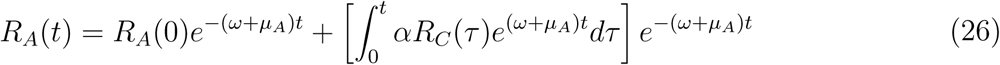

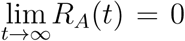 Solving for *R*_*A*_(*t*) and substituting the value of *S*_*A*_(*t*) in the expression obtained gives

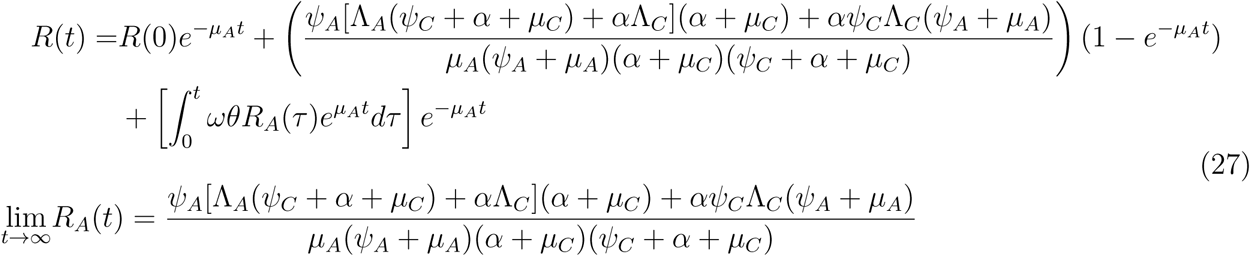

Solving for *S*_*C*_ (*t*) gives

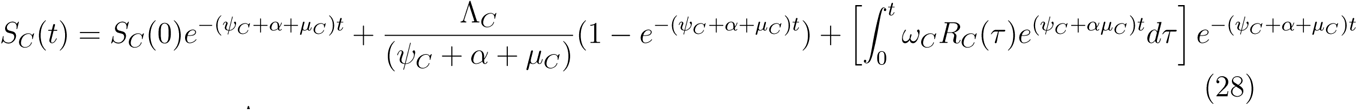

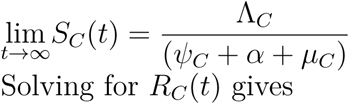

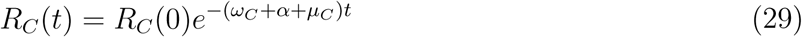

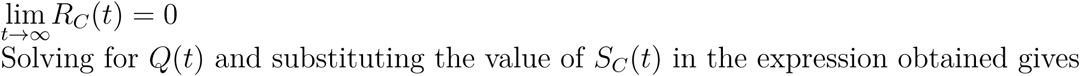

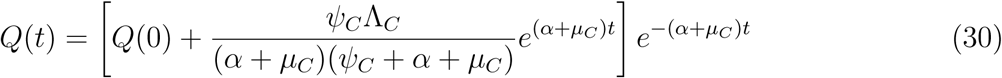

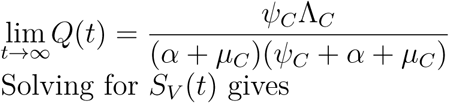

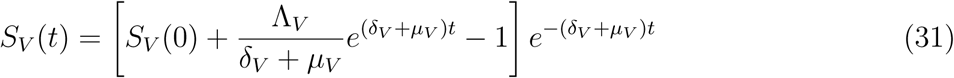

taking limit of S_*V*_ (t) as t → ∞ givesV

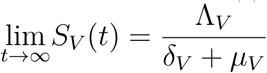

The asymptotic dynamics are independent of initial conditions in *D*. Hence, the solutions of (23) converge globally in *D*. According to previous study [16] it is required to show that *G*(*X, Z*) satisfies the two stated conditions

i. *G*(*X*, 0) = 0 and
ii. *G*(*X, Z*) = *D*_*z*_*G*(*X*^*\**^, 0)*Z* − *Ĝ* (*X, Z*), *Ĝ* (*X, Z*) ≥ 0 where

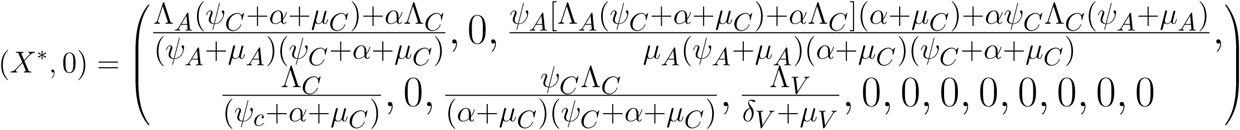

*D*_*z*_*G*(*X*^*\**^, 0) is the Jacobian of *G*(*X, Z*) taken with respect to the infected classes and evaluated at (*X*^*\**^, 0).

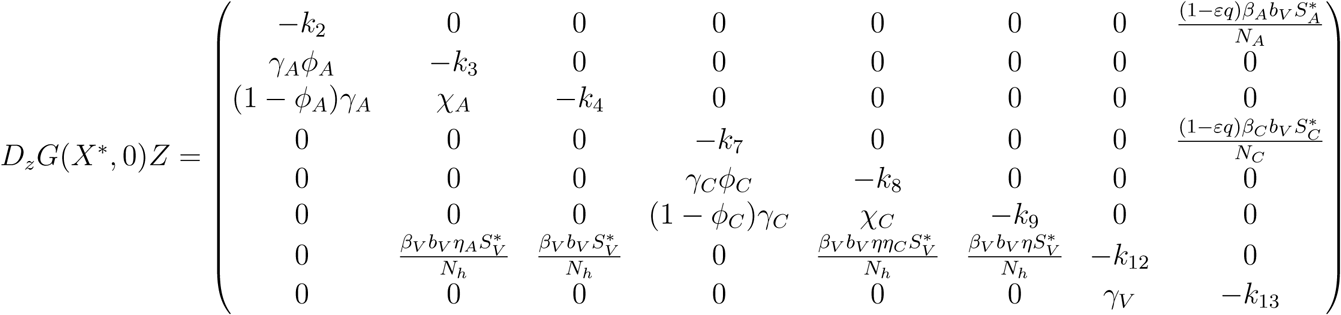

and

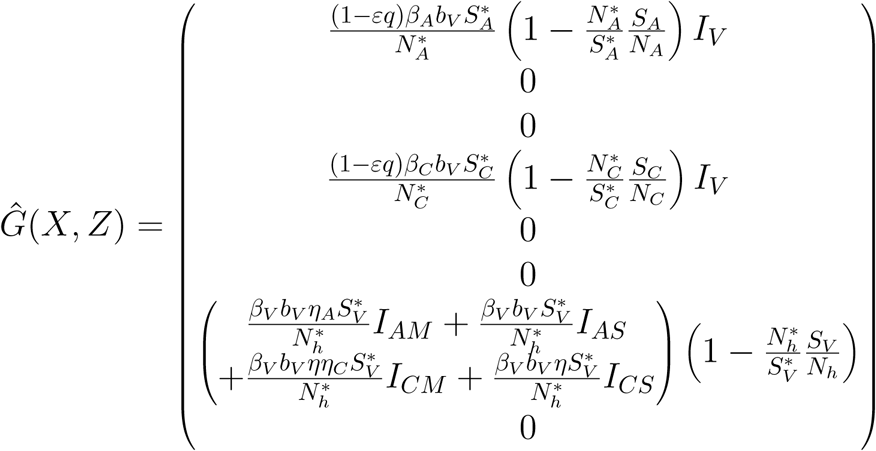

Since we have

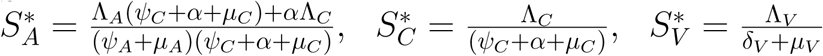

In *D*, 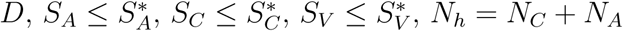 and thus 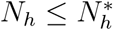

If the human population is at equilibrium, we have

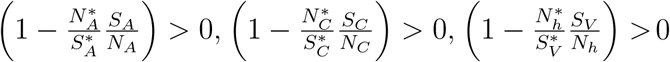 ; thus *Ĝ* (*X, Z*) ≥ 0. Therefore, the DFE is globally asymptotically stable by the theorem in [16].

***Proof:***(Proof of Lemma 7)

Suppose *ℛ*_*E*_ > 1 then the existence of the endemic equilibrium point is guaranteed. Using the common quadratic Lyapunov function,

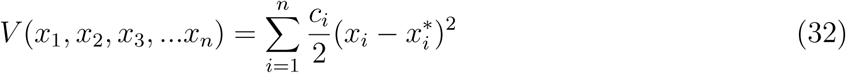

As formerly illustrated [20], we consider a Lyapunov function with the following state variables

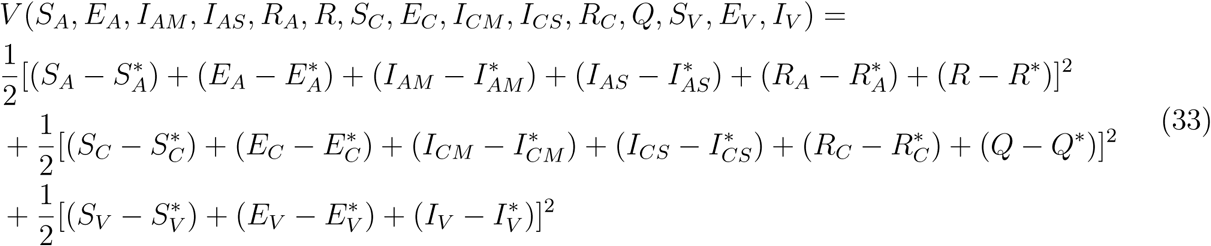

Now, differentiating (33) along the solution curve of (32) gives

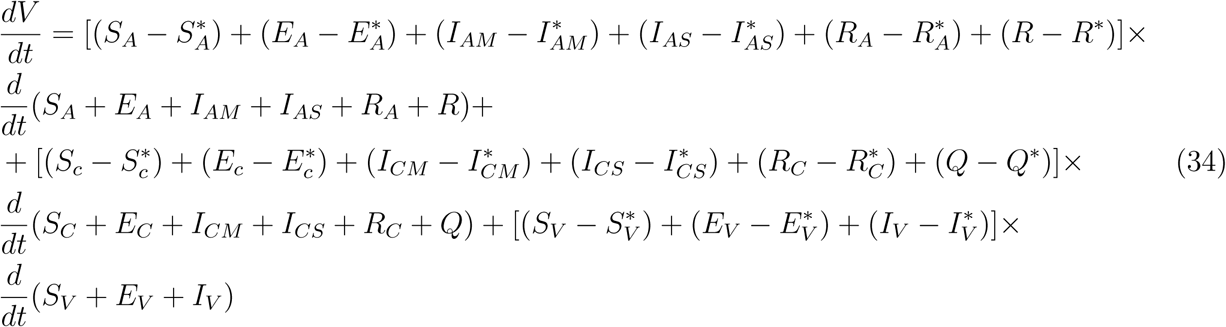

from (32), it implies that

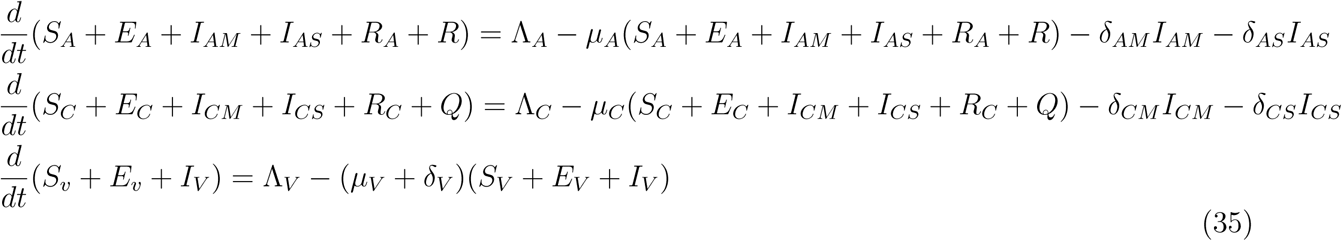

Plugging (34) to (35) gives

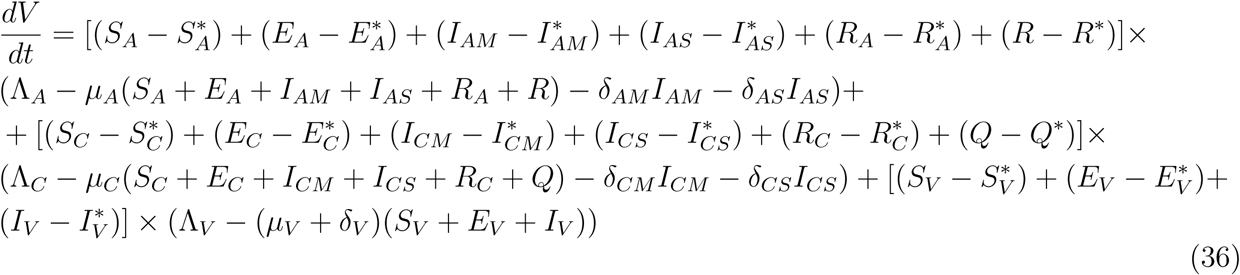

Now assuming,

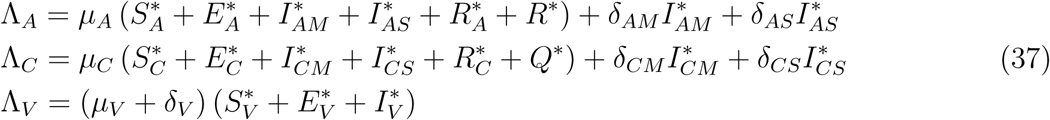

Substituting (37) into (36)

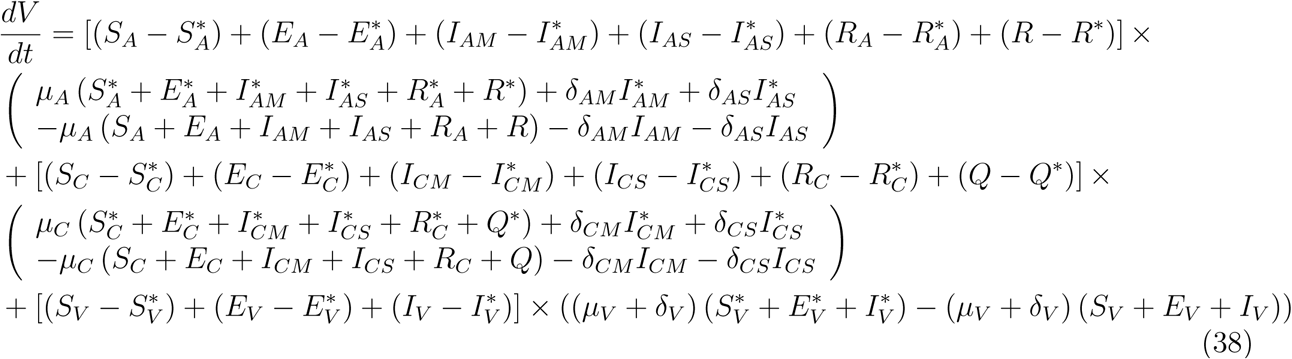

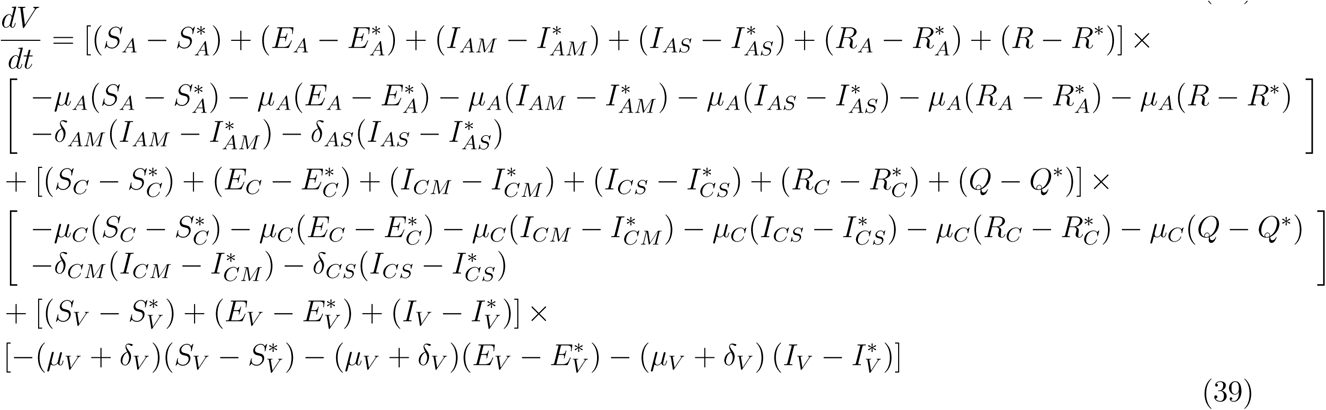

This implies that

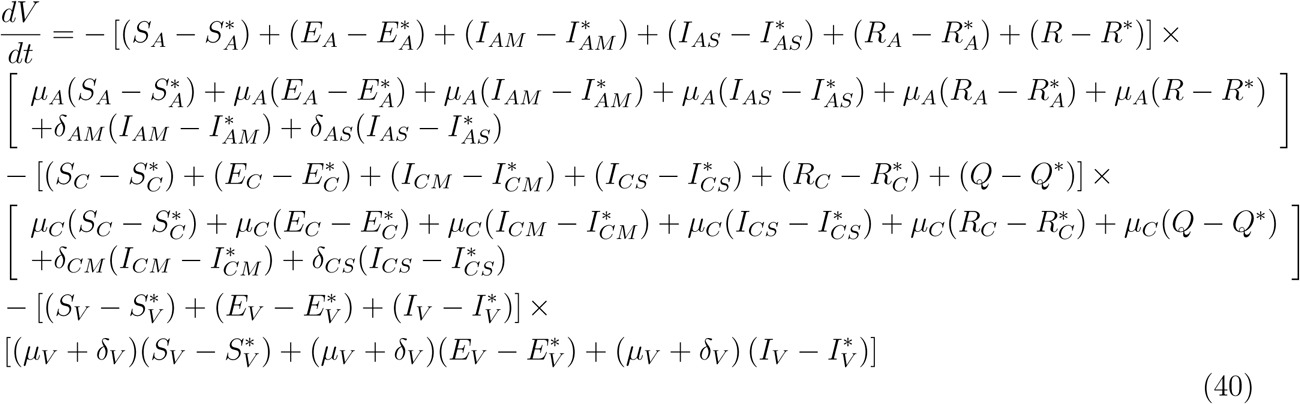

This shows that 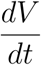 is negative and 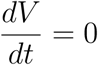 if and only if

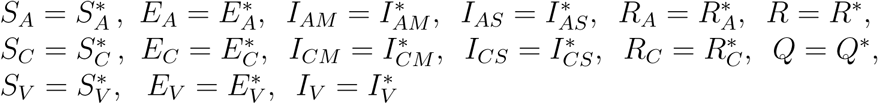

Additionally, every solution of (1) with the initial conditions approaches *ξ*^*\*\**^ as *t* → ∞.

Therefore, the largest compact invariant set in

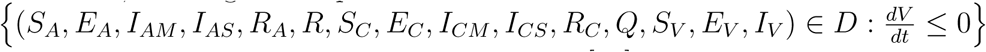 is a singleton set {*ξ*^*\*\**^}

Therefore, from Lassalle’s invariant principle [21], it implies that the endemic equilibrium *ξ*^*\*\**^ is globally asymptotically stable in D whenever *ℛ*_*E*_ > 1

